# Adverse mental health inpatient experiences: Qualitative systematic review of international literature and development of a conceptual framework

**DOI:** 10.1101/2023.10.20.23297217

**Authors:** Nutmeg Hallett, Rachel Dickinson, Emachi Eneje, Geoffrey L. Dickens

## Abstract

**Background:** Trauma is increasingly linked to poor health outcomes. Adverse experiences in mental health inpatient settings can be traumatic and contribute to long-lasting negative effects like post-traumatic stress disorder. However, the full range of relevant experiences is often unaddressed in service design and delivery.

**Aim:** To describe the spectrum of negative experiences that people identify while they are inpatients in adult mental health services.

**Method:** A systematic literature review was conducted to identify qualitative studies that included people’s subjective negative reports of their inpatient admissions. CINAHL, MEDLINE and PsycINFO were searched from 2000 onwards, alongside a search of Google Scholar. The quality of studies was assessed using the Critical Appraisal Skills Programme qualitative checklist. Data were synthesised using the ‘best-fit’ framework synthesis approach. A patient and public involvement reference group contributed to the review.

**Results:** Studies (111) from 25 countries were included. Adverse mental health inpatient experiences can be conceptualised under three headings: the ecosystem (the physical environment and the resources available, and other people within or influential to that environment); systems (processes and transitions); and the individual (encroachments on autonomy and traumatisation).

**Conclusions:** Improved patient experience is associated with improved patient outcomes, and addressing negative experiences could significantly impact patient care. Mental health professionals should strive to create inpatient environments that are supportive, respectful, and safe for patients, which consideration of the adversity framework developed from this review can facilitate.

## Introduction

There is increasing evidence of links between trauma and subsequent ill health. The frequency and severity of adverse childhood experiences (ACEs) are reportedly linked to negative mental ill health in later life in a dose-response relationship.^1^ The relationship between mental ill health and ACEs has been demonstrated globally.^2^ For already vulnerable adult populations, such as those with psychosis or other serious mental health conditions, adverse experiences in mental health inpatient settings may be related to, for example, Post Traumatic Stress Disorder.^3^ The most restrictive interventions in mental health inpatient settings, namely seclusion, restraint and rapid tranquillisation, are commonly practised with differing frequency and varying acceptability throughout the world and are recognised sources of potential harm to patients; estimates of PTSD following restrictive interventions range from 25% to 47%.^4^

While the restrictive interventions described above can have clear adverse effects, other experiences in the inpatient mental health setting can also be negative. A review of patients’ perceptions of antecedents and consequences of coercion found that being subjected to professionals’ control was an important negatively experienced precursor of coercion, while communication and interactions with professionals were notable along the timeline of coercion.^5^ Staniszewska et al,^6^ in a broad review of inpatient mental health experiences, identified ward milieu, boredom and lack of information as negative experiences alongside restrictive interventions. To our knowledge, no previous review has set out to identify and synthesise experiences that inpatients describe negatively either in themselves or their subsequent impact. This is important if we are to better understand inpatient admissions from the perspective of the people who experience them; improved patient experience is associated with better outcomes.^7^ It is imperative that services have the willingness to examine where they fail patients because if they do not, how will they know where improvements can be made? Capturing the patient’s voice is an important first step in achieving this. The aim of this systematic review, therefore, was to identify the spectrum of negative experiences that people describe while they are inpatients in adult mental health services.

## Method

The protocol for this review was registered with the International Prospective Register of Systematic Reviews (PROSPERO; CRD42022323237) and can be accessed at https://www.crd.york.ac.uk/prospero/display_record.php?RecordID=323237. This review is reported according to the Preferred Reporting Items for Systematic Reviews and Meta-Analyses (PRISMA) guidelines.^8^

### Search strategy

The search strategy was developed using the Sample (inpatient and psychiatry), Phenomenon of Interest (harm/adversity), Design (interview, focus group, questionnaire), Evaluation (experience/satisfaction), Research type (qualitative) (SPIDER) framework.

Search terms were developed based on the sample, phenomenon of interest and evaluation, and applied in CINAHL, MEDLINE and PsycINFO searches. Medical Subject Headings (MeSH) were used where available, see Supplementary Table 1. Searches were limited from 2000 onwards to ensure contemporary literature only and to reports in English. Supplementary searches were conducted in Google Scholar; 200 results were extracted using Harzing’s Publish or Perish software. Results were exported into Rayyan^9^ for de-duplication and screening. The search strategy deviated from the protocol in that we did not conduct forward and backward chain searching of included studies due to the large number of included studies.

### Eligibility criteria

Qualitative studies, to ensure that the patient voice is captured, conducted with current or former inpatients of adult acute, forensic and psychiatric intensive care services were included. Studies in community mental health and general hospital settings were excluded, as were studies in specialist settings such as child and adolescent mental health, older adults and learning disabilities settings. Studies had to include participants’ subjective reports of their inpatient experiences that were described in negative terms; we included studies that looked at admission and discharge experiences but excluded studies that examined experiences pre-or post-discharge, for example, community treatment orders.

### Study selection

Due to the number of hits, study titles and abstracts were screened by a single reviewer (NH) and 10% were independently screened by a second reviewer (RD). Full texts of studies retained after screening were independently assessed for eligibility against the criteria by two reviewers (NH, RD).

### Data extraction

Data were extracted by a single reviewer (RD) using a bespoke data extraction table: authors, year, country, aim, study design, data collection, time of data collection, setting, participants, and authors’ themes/results with illustrative quotes. Extracted data were checked by a second reviewer (NH).

### Quality and risk of bias

The 10-point Critical Appraisal Skills Programme qualitative checklist was used to assess study quality.^10^ Studies were not excluded based on quality; quality assessment was used to explore the robustness of our synthesis.^11^

### Data synthesis

We synthesised the data using the “best fit” framework synthesis approach.^12^ The a priori framework was developed through examination of the themes identified in the review by Staniszewska et al^6^ and from discussions between the authors around which themes could be described as adverse. The initial framework comprised: seclusion, restraint, forced medication/sedation, involuntary admission, police involvement, lack of choice and separation from family. This framework guided but did not restrict the review. Data that did not fit the framework were extracted and analysed thematically and the framework was adapted accordingly as analysis progressed.

### Patient and public involvement reference group

PPI is detailed in the Guidance for Reporting Involvement of Patients and the Public 2^nd^ version (GRIPP2) form (Table 1).^13^ The Patient and Public Involvement Reference Group (PPIRG) included five service users with varied experiences of acute and secure inpatient care. After analysis of around half the included studies, we presented the preliminary framework of findings to the PPIERG. The PPI author has been involved at all stages of the review (EE).

**Table 1.**
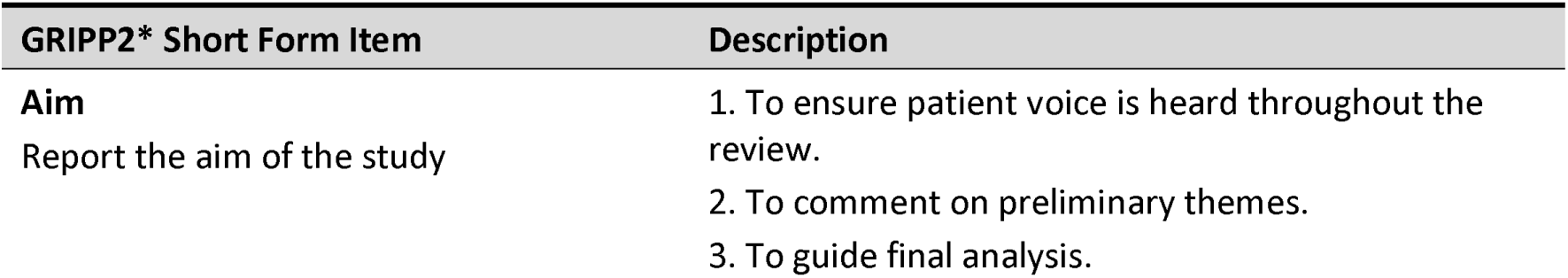
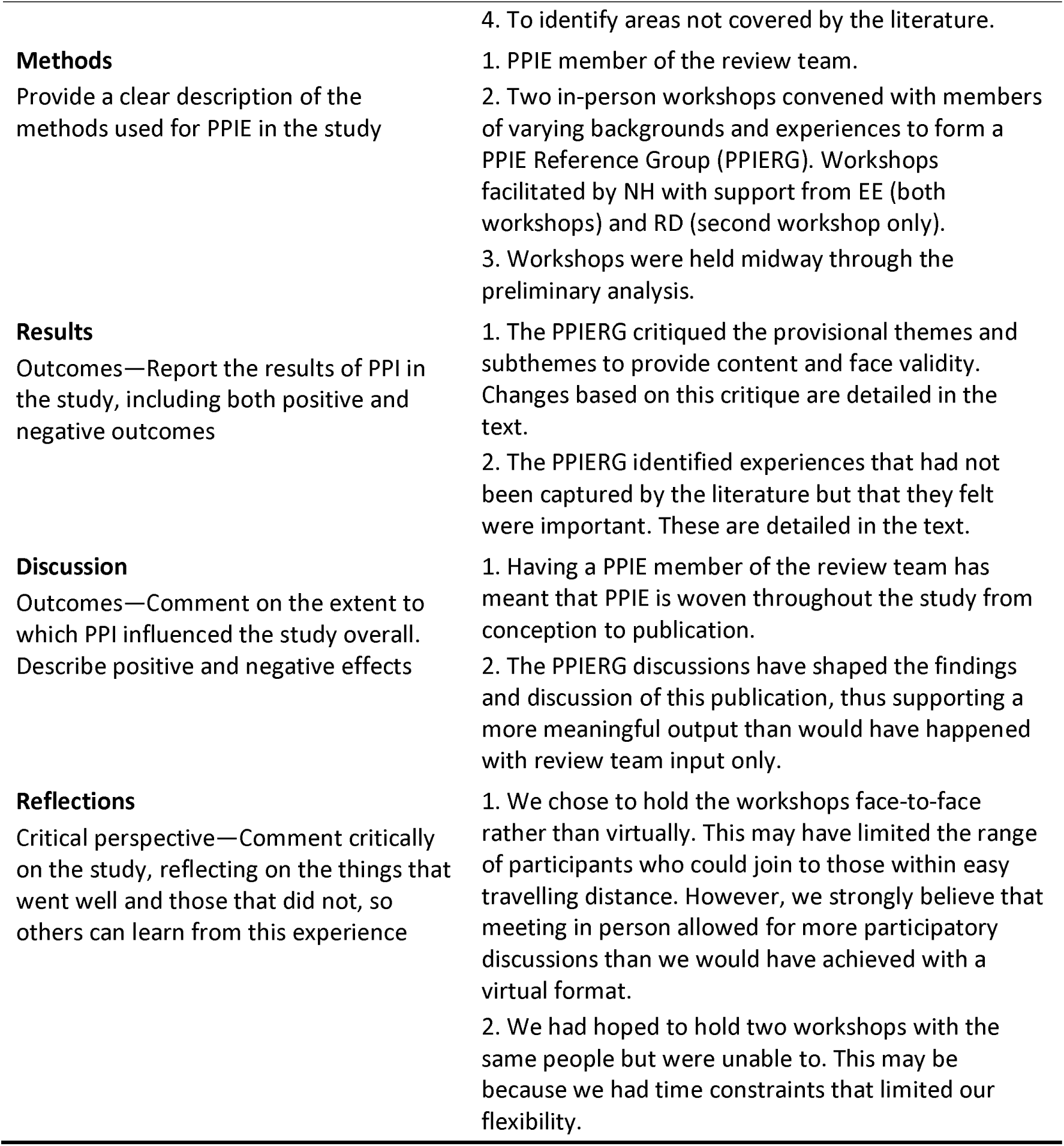
Public and patient involvement.

## Results

After removal of 782 duplicates from the 4,012 records identified, 3,230 were screened by title and abstract, resulting in 3,046 exclusions (see Figure 1). We were unable to access five records so 179 full texts were assessed against the eligibility criteria resulting in 111 papers included in this review.

**Figure 1.**
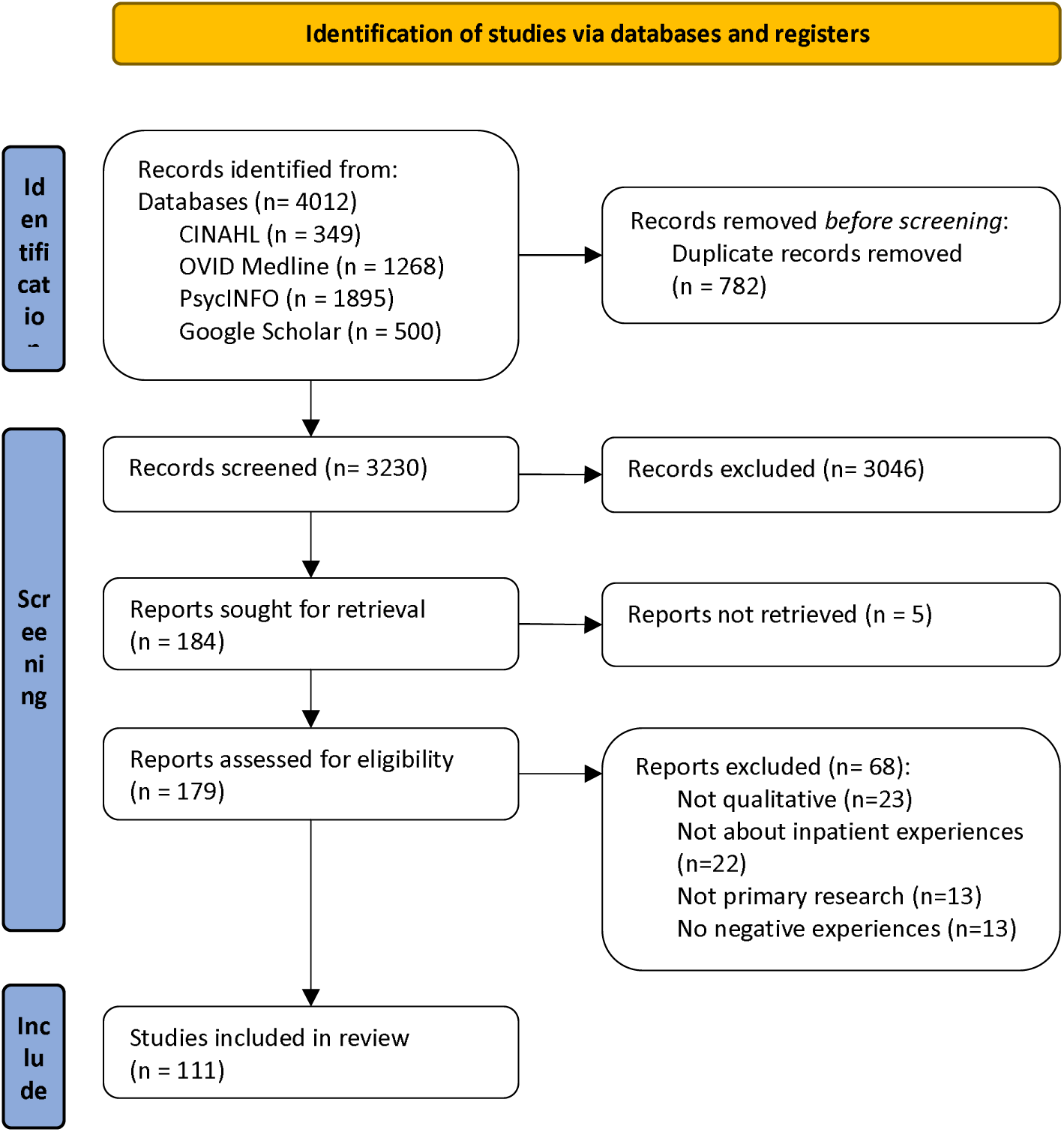
*PRISMA flow diagram from: Page MJ, McKenzie JE, Bossuyt PM, Boutron I, Hoffmann TC, Mulrow CD, et al. The PRISMA 2020 statement: an updated guideline for reporting systematic reviews. BMJ. 2021; 372: n71.*

### Study characteristics and quality

The studies spanned the globe but were predominantly conducted in Europe (n=80), see Supplementary Table 2. Four were conducted across multiple European countries. The UK was noticeably over-represented (n=42). Other study locations were Sweden (n=12), Canada and USA (n=9 each), Norway (n=7), Australia (n=6), Ireland (n=4), Finland (n=3), Belgium (n=2), Germany (n=2), and one each in Austria, Brazil, Hong Kong, Israel, Italy, Japan, Lesotho, New Zealand, Spain, South Africa and The Netherlands.

The sample size of the studies ranged from 1, an autoethnographic study,^14^ to 906, taken from the qualitative responses to a service user care experience survey,^15^ with a median (IQR) sample size of 10 (20). In total there were 4,255 participants; one study recorded patient observation hours but not the number of patients observed.^16^

Many studies explored experiences of hospitalisation (n=22) or coercion in hospital (n=9), whilst others focused on specific aspects or types of hospitalisations: involuntary admissions (n=6) and conversely, voluntary patients (n=1), and forensic (n=5), or specific phenomena, namely seclusion (n=10), restraint (n=7), and seclusion and restraint (n=2). Some studies focused on specific populations: women (n=5), people who self-harm (n=3) and people with a diagnosis of anorexia nervosa (n=2), schizophrenia (n=2), autistic spectrum disorder (n=1), depressive psychosis (n=1), eating disorder (n=1), intellectual disabilities (n=1) and personality disorder (n=1).

The studies largely collected data using semi-structured interviews (n=69), see Supplementary Table 2. Other methods used were alternative interview methods (n=10), focus groups (n=7), surveys or questionnaires (n=5) or a combination of interviews, focus groups and/or qualitative surveys (n=12). Data collection in most studies was conducted while participants were still inpatients (n=53); in the remaining studies it was conducted following discharge (n=35) or at a combination of time points (n=12). Eleven papers did not report the time point of data collection.

The research was variable in quality, with many of the papers being identified as poor to medium quality according to the CASP checklist.

### PPIERG input

There was broad agreement with the framework. However, group members said that ‘trauma/retraumatisation’, which we had captured as a subtheme related to practices including seclusion and restraint pervaded the whole inpatient experience and so should be captured as a theme in its own right. Group members expressed that a thematic element preliminary named ‘loss of power’ was insufficiently nuanced and suggested instead that it was an imbalance of power between staff and patients that more accurately captured the experience. The PPIERG also said that a lack of reference to racism and cultural needs was a notable omission in the framework. In response, and before analysing the remaining papers, we adapted the framework to include trauma as a theme and revisited studies to capture issues about power imbalances, and racism. We remained mindful of power and racism in analyses going forward.

### Findings

Adverse mental health inpatient experiences can be described under three main headings, each comprising two linked themes. ‘The ecosystem’ describes i) the physical environment in which adverse experiences occur, most notably the ward and the hospital; also the resources available to that environment and ii) other people within or influential to that environment with whom interactions that can be experienced as adverse occur. ‘Systems’ describes the formal mechanisms of psychiatric treatment that can be experienced as adverse; we have separated these into i) processes – tangible formal practices of professionals that can be experienced as adverse, and ii) transitions – discrete periods in which a change of circumstances or environment occurs. Finally, ‘the individual’ describes i) the encroachments on autonomy that are perceived as adverse; and ii) the traumatisation and/or retraumatisation that might be attributable in part or whole to the totality of adverse experiences.

### The ecosystem

#### Physical environment

The physical environment of, and tangible resources available in, the inpatient setting were described negatively. A dirty or poor ward environment was perceived as an adverse experience exacerbated by other ward experiences.^17–19^ Overbearing sensory stimuli, including bright hospital lighting, the strong smell of cleaning products, the taste, smell and texture of hospital food, and the cacophony of heavily closing doors, buzzer systems, and intermittent beeping of fire alarms, were referred to negatively, particularly for neurodiverse respondents. The combination of stimuli in the physical environment was likened by participants to ‘sensory overload’.^20, 21^.

A lack of activities, therapies or resources was frequently recounted. This seemed to reflect a gap between expectation and reality.^22–27^ Resulting feelings of boredom were often cited as directly worsening mood and mental state.^19, 24, 28–36^ Some papers reported that, while activities or groups were available, they were not delivered in a person-centred manner that met participants’ needs resulting in non-engagement [44]. When access to coping strategies was removed, which was sometimes perceived as arbitrary, participants felt unheard by staff. Across the literature, there were reports that the rules and boundaries in inpatient settings were unfair, overly restrictive, and inconsistently implemented.^16, 17, 21, 30, 31, 34–46^ This inconsistency in rules led to perceptions of staff to patient favouritism, persecution and punishment.

Hospital wards were frequently spoken of as prison-like environments.^14, 17, 19, 27, 30, 31, 34–36, 38–42, 47–61^ This was exacerbated when staff were perceived as custodial and lacking in caring or nurturing qualities.^17, 36, 39, 40, 54, 56^ Integral to the perceived custodial environment were the set routines around mealtimes and bedtimes which were viewed as infantilising, and as leading to loss of identity and individuality.^21, 36, 55, 61, 62^ Night times were raised as problematic because participants in some studies reported feeling unsafe at night.^19, 33, 38, 60, 63^ Close and intermittent observations throughout the night were seen as highly disruptive due to staff turning on lights or using torches at intervals.^20, 33, 38,60, 61, 64, 65^ Wards were also described as loud and not conducive to rest.

#### Other people

The second part of the ecosystem encompassed other people with whom participants had interactions during their inpatient stay, usually the ward staff, but also fellow patients and family members. Lack of staff availability was viewed negatively by some patients and was believed to be due to low staffing ratios,^36, 46, 58, 66–68^ poor staff visibility,^17, 24, 33, 46^ an office-based culture^24, 35, 46,48, 58^ and, perceived social distancing behaviour by staff ^18, 25, 26, 29, 30, 46, 58, 69^

Poor staff communication was discussed in many studies. Frequent use of inaccessible language and jargon was a barrier to effective communication.^53, 62, 70^ Participants reported a lack of information-sharing by staff.^16, 18, 25, 26, 33, 46, 55, 61, 62, 68, 71–74^ Lack of involvement in decision-making processes was viewed as highly distressing, impacting personal autonomy and inherently limiting the therapeutic nature of inpatient care.^18, 19, 24, 27, 30, 33, 36, 46, 49, 57, 68, 71, 75–77^ Extrapolating from this, adverse experience emerged from an incongruence between the expectations or beliefs of the staff and those of patients,^32, 36, 50, 78–82^ a phenomenon described as a battle of expectation versus disappointment. Patients said they wanted to feel cared for, yet instead felt punished or dehumanised by staff; this was perceived as an injustice, and left patients feeling their views were unimportant or disregarded.^32, 50, 80, 81^ This incongruence was also described by those in forensic services: participants described the restrictions as feeling more salient when they perceived a difference between what they expected and what they lived with.^36^

Staff attitudes were commonly described as unprofessional, uncaring and unfair.^15, 17, 19, 21, 24, 26–28, 31, 32,35, 37, 38, 40, 46, 48–50, 52, 53, 59, 62, 66, 68, 72, 83–95^ This reportedly contributed to a negative ward atmosphere and culture, making the overall admission experience poorer. Participants in many studies described feeling infantilised and patronised by staff; this was explained as arising in the context of a parent-child or teacher-child relationship, where one party has significantly less power and control, leading to feelings of disrespect;^24, 27, 38, 42, 44, 72, 73, 85, 92, 94, 96^. Similarly, staff behaviour was frequently described as dehumanising.^17, 21, 24, 27, 32, 34, 36, 37, 42, 46, 48, 59, 62, 68, 87, 97^

Patients in many studies said they felt hesitant to express their emotions, attitudes, values or beliefs during their inpatient stay due to a fear of them being used by staff as evidence of psychopathology. Participants reported that their beliefs, if different from those in mainstream society, were viewed as symptoms of mental illness.^24, 25, 27, 30, 31, 37, 43, 44, 67, 72, 73, 91, 92^ Some participants felt unable to express their religious beliefs as they had witnessed others who did so being labelled as displaying religiose delusions.^27, 73^ Others described concealing their emotions, particularly anger, out of fear of it being problematized as potentially aggressive and thereby increasing the risk of restraint and seclusion.^25,31, 37, 44, 72, 91, 92^

Issues of racism and cultural insensitivity were infrequently discussed. Predominantly, participants perceived being treated less favourably because of their ethnicity.^26, 48^ Several described being viewed as dangerous because they were black, reporting that staff were quicker to restrain, medicate or seclude them than their white counterparts.^26, 44, 48^. A lack of cultural awareness was described by some participants with staff either stereotyping patients based on their ethnicity or lacking understanding about patients’ beliefs or needs.^26, 48^

In some papers participants said they had been stigmatised by staff: patients described being labelled as bad or attention-seeking. This was mainly reported by patients with personality disorder^32, 49^ and substance misuse diagnoses^50^ but also by those in forensic services and patients who self-harm.^32, 35^ This perceived stigma was experienced predominantly as a judgment and withdrawal of caring from staff.

Participants frequently referenced vicarious exposure to seriously mentally disordered patients, sometimes in distressing situations, particularly those involving coercive practices.^25, 27, 38, 62, 71, 75, 89, 91,98^ This culminated in negative feelings towards both the patient and staff members involved.^33, 34, 36, 71, 95, 99, 100^ The fear of threatened or actual assault from fellow patients was relatively infrequently reported but remained a salient factor in perceived adverse experiences.^17–19, 36, 48, 60, 62, 79, 89, 91, 98, 101,102^

Participants also experienced adversity arising from their family relationships, largely from an enforced lack of interaction. During admission, participants described the lack of choice over the location of the unit and were often geographically distanced from family, friends, community and culture.^15, 36, 62, 71^ This separation was worsened when family were uninvolved in participants’ care^15,17, 30^ and information was not shared appropriately with family.^30, 67^ Several papers reported a lack of a family-centred approach, i.e. considering the parent, child and family as a whole, with little regard given to those separated from their children during admission.^19, 31, 102^ Finally, the impact of admission and mental health deterioration was linked to worsening relationships with family and friends for some participants.^36, 67, 102, 103^ They reported that being detained led to fear of stigma from family and friends, a factor which some continued to report following discharge.^54, 72^

### Systems

#### Processes

Processes encompass exposure to the application of formal aspects of mental health treatment including coercive management strategies, the use of psychotropic medication, the legal process in the form of mental health tribunals, and the monitoring of progress in the form of ward rounds.

Seclusion and restraint were described as highly distressing experiences and participants who had previously been subjected to these practices reported avoiding hospital admission due to fear of further episodes.^57, 71, 84, 104^ This was described as leading to a further deterioration of mental state, and increased risk of the use of these measures when eventually admitted.^17, 27, 35, 43, 44, 57, 71, 88, 105^ Despite this belief, the fear continued to act as a barrier to service access.

There was extensive reference to a fear of staff-perpetrated violence during restraint amounting to the use of excessive force or in the use and duration of seclusion.^21, 39, 40, 43, 48, 49, 63, 71, 73, 81, 97, 99, 104, 106–108^ It is difficult to establish from the literature any objective threshold whose transgression could be deemed as excessive. However, there was a sense from participants that staff were to be feared and that injury was a likely outcome of restraint. Indeed, many participants reported physical injury from restraint.^28, 38, 48, 63, 81, 104^ In several studies participants reported that witnessing the restraint or seclusion of peers was a traumatic experience in its own right.^33, 34, 36, 95, 99, 100^ Fear, shame and humiliation were visceral reactions to seclusion and restraint.^39, 42, 43, 47, 49, 84, 95, 97, 99, 100, 104, 109–112^ The experience was variously described as punitive,^39, 47, 71, 80-82, 95, 99, 100, 104, 107, 108, 110, 112-114^ untherapeutic,^18, 39, 45, 50, 81, 82, 97, 100, 104, 107, 111, 112^ dehumanizing^21, 28, 39, 42, 47, 63, 71, 81, 82, 99, 100, 106, 108-111, 114^ and traumatising.^39, 40, 47, 57, 81, 82, 84, 88, 104, 109, 112, 114^ Factors which were reported to impact the experience of restraint and seclusion included poor communication from the staff involved^57, 63, 84, 108^ and dearth of appropriate debriefs following these incidents.^71, 82, 84^ Seclusion and restraint were commonly considered to worsen overall treatment compliance and therapeutic relationships with staff.^18, 39, 45, 50, 81, 82, 97, 100, 104, 107, 111, 112^

Psychotropic medication was discussed in most studies, predominantly negatively. Participants reported an overreliance by practitioners on medication and believed that decisions were largely informed by a medical model of mental distress, which was regarded by participants as lacking person-centredness.^14, 16, 17, 19, 24-27, 35, 57-60, 71, 72, 76, 90, 96, 97, 102, 103, 105^ Participants reported that medication was the first line of treatment and that there was a lack of talking, holistic or person-centred approaches offered before its use. Many felt coerced into taking medication;^14, 16, 28, 36–38, 43, 44,48, 49, 58, 72, 73, 89, 103^ participants were not given choices about whether to take medication but were threatened with restraint, seclusion, forced injection or involuntary detention should they decline.^17,30, 48, 72^ Participants frequently considered forced medication to be punitive.^25, 34, 37, 39, 43, 73, 91, 102, 106^ It was reported across several papers that forced medication was used in response to perceived bad behaviour and as a way to control patients.

Mental Health Tribunals were less infrequently reported as adverse experiences. Tribunals are an important point of care during involuntary admissions and participants cited a lack of support around their occurrence.^57, 115^ Many participants discussed poor communication from staff, feeling unprepared, and being unsure of what to expect in the tribunal.^30, 57, 61, 115^ During the tribunal, several studies reported a power imbalance between the patient and the professionals on the tribunal panel.^30, 57, 61, 115, 116^ Some studies noted that the tribunal process appeared to participants as a formality with only one possible outcome, namely continued detention.^61, 115, 116^ Ward rounds were, as tribunals, described rarely but were viewed as intimidating and unhelpful.^77, 79^ This was reported in reference to power imbalance, the feeling of having no voice and poor communication between staff and patient.

#### Transitions

‘Transitions’ describes aspects of treatment involving exposure to transition resulting from progress, or indeed regress, through the mental health system; this includes admission, transfer, and discharge.

Admission to hospital, voluntarily or involuntarily, was widely associated with fear.^19, 22, 24, 30, 46, 48, 51, 57,59, 61, 75, 88, 102, 106, 117^ Participants repeatedly stated that a voluntary admission was, in reality, the result of coercion and threats.^30, 35, 42, 44, 45, 48, 62, 73, 117^ Many participants reported being admitted via the police or secure transport, which contributed to feelings of fear and shame,^25, 51, 57, 72, 73, 105, 117, 118^ and, particularly for those admitted from home with friends or family witnesses, stigma.^105, 118^ Police involvement was also experienced as punitive and likened to a custodial rather than a healthcare experience.^51, 73, 105, 117, 118^ Those detained under mental health legislation reported that this in itself was stigmatising; participants described involuntary detention as synonymous with dangerousness, a factor which was exacerbated by police involvement.^25, 73, 119^

Transfer between wards, or between different services, was less prominent in the literature but provided a negative experience for some. Participants felt unprepared for transfer due to a lack of clear communication about the process.^15, 18, 57, 74, 98, 116, 120^ Decisions about transfer were viewed as processes that happen to rather than with patients. ^18, 98, 116, 120^ This could lead to a heightened sense of loss of control.

Similarly, feelings of unpreparedness due to lack of involvement in planning for discharge were reported.^17, 35, 46, 50, 67, 75, 78, 91, 116, 120^ There were many reports of insufficient notice before discharge, contributing to this lack of preparedness on emotional and practical levels.^17, 46, 50, 78, 120^ One study reported that participants were discharged prematurely due to insurance not covering treatment costs or having met the maximum amount allowed.^34^ For these participants, this perpetuated the cycle of avoiding admission due to costs and fear of poor treatment.

### Individual

#### Autonomy

Patients’ autonomy, or specifically the threats to it, seemed central to the adverse patient experience and included issues of perceived loss of control, privacy and freedom, power and choice.

Loss of control was predominantly spoken about in relation to coercive practices and the resultant feeling of voicelessness.^15-19, 33, 36, 37, 43, 46, 48, 50, 55, 57, 59, 61, 62, 73, 84, 90, 93, 95, 102, 103, 105, 106, 108, 109, 112^ The sense of loss of control over one’s choices about admission and treatment led to despondency and worsened the overall view of other aspects of admission. Wards were commonly described as having overly restrictive rules about personal possessions, clothing and money.^16, 17, 21, 31, 35, 36, 38–41, 43^ This was felt to be controlling and coercive and to limit patients’ autonomy. Several papers referred to restrictions on behaviour on the ward to the extent that limits were placed on recognised coping strategies, such as smoking cigarettes, listening to music and maintaining routines.^31, 38, 40^

Loss of control was also experienced in the rigidity of ward routines including set mealtimes and bedtimes ^21, 36, 55, 61, 62^ and was expressed as infantilising.^24, 38, 42, 44, 72, 73, 80, 92, 94, 96^ Loss of privacy was also reported, especially in relation to enhanced observations,^14, 23, 27, 30, 31, 34, 35, 37, 41, 46, 48, 52, 54, 57-59, 62, 72, 80, 86, 94, 95, 105, 112^ which were experienced as intrusive, infantilising and non-therapeutic.^17, 27, 49, 64, 65, 83, 87, 93^ Loss of freedom was described in terms of confinement and containment. Locked doors were commonly reported as physically confining as well as a visual reminder of their loss of freedom,^31, 48,54, 57, 62, 90^ a feeling exacerbated by limitations on hospital leave due to having insufficient staff to facilitate escorts.^36, 41, 43, 94^ Informal patients also experience this loss of liberty and report restrictions placed upon their freedom to leave.^30, 35, 48^

Descriptions of the loss of power and power imbalances were common. Many studies reported that admission and detention under mental health legislation were inherently associated with loss of power.^14, 15, 24, 30, 34, 37, 41, 46, 47, 49, 50, 52, 55, 67, 75, 79, 86, 88, 99, 100, 106, 109, 121^ The relationship between staff and patients was recognised as having an innate power imbalance, regardless of the characteristics of the individuals involved.^18, 30, 34, 35, 39, 52, 79, 86, 95, 97, 99, 100, 104, 106, 109, 113, 116^ Two papers also referenced a perceived power imbalance between patients on the ward,^24, 103^ though this appeared less impactful on autonomy.

Feelings of restricted choice due to involuntary admission and forced treatment were regularly linked to a perceived loss of autonomy.^24, 25, 27, 43, 57, 59, 68, 73, 74, 76, 90, 95^ However, of apparent greater concern was the issue of false choices.^73, 116^ A pertinent example concerned voluntary patients who frequently reported that, despite the non-enforced status of their admission, they were given the choice of being admitted to or remaining in hospital voluntarily or being detained under mental health legislation.^42, 44, 45, 48, 62, 69^ Many of the same participants reported similar non-choices, such as taking medication voluntarily or being compelled to do so.^17, 30, 35, 48, 72, 73^

#### Trauma and retraumatisation

Finally, trauma and retraumatisation were evident throughout the literature. Both restraint and seclusion were described as traumatic experiences, regardless of previous trauma history.^15, 39, 40, 47, 52, 81, 82, 84, 86, 88, 104, 109, 112, 114, 121^ Patients reported trauma responses to previous experiences of restraint and seclusion. Those who spoke of retraumatisation primarily referred to histories of sexual or physical abuse, which they reported leading to heightened trauma responses leading up to, during and following incidents of seclusion or restraint.^39, 40, 81, 82, 86, 102, 104^ Frequently these experiences led to flashbacks and feelings of humiliation. Mixed-gender wards were described as difficult environments for those with histories of physical and sexual abuse, whether or not an incident took place on the ward; the mixed-gender ward itself was perceived as triggering and as an adverse experience.^17, 36, 79^ Racism and trauma were intertwined within the literature. Racial inequalities were often described in relation to previous experiences of racism.^26, 44,48^ Racially based traumas were reported in the context of interpersonal racism from staff and systemic, institutional racism.

## Discussion

To our knowledge, this is the first review to attempt to identify and synthesise literature from primary research about the negative experiences that are described by mental health inpatients. While other reviews and individual studies provide snippets of such experiences, this is the first time that the full range of negative experiences, as described in the literature at least, have been laid bare. The review identified three overarching themes, namely: the ecosystem, systems and the person. These themes can assist a comprehensive understanding of the spectrum experiences that patients encounter during their stay in mental health inpatient settings and perceive to be negative A previous review^6^ explored themes for improving inpatient experiences, which formed the basis for our “best fit” framework. Each of the factors identified in this preliminary framework was evident in the results, however, a far wider range of negative experiences was revealed. There was some consensus on what constituted negative experiences across the literature, despite different ward environments, sample participants and countries of study.

**Figure 2.**
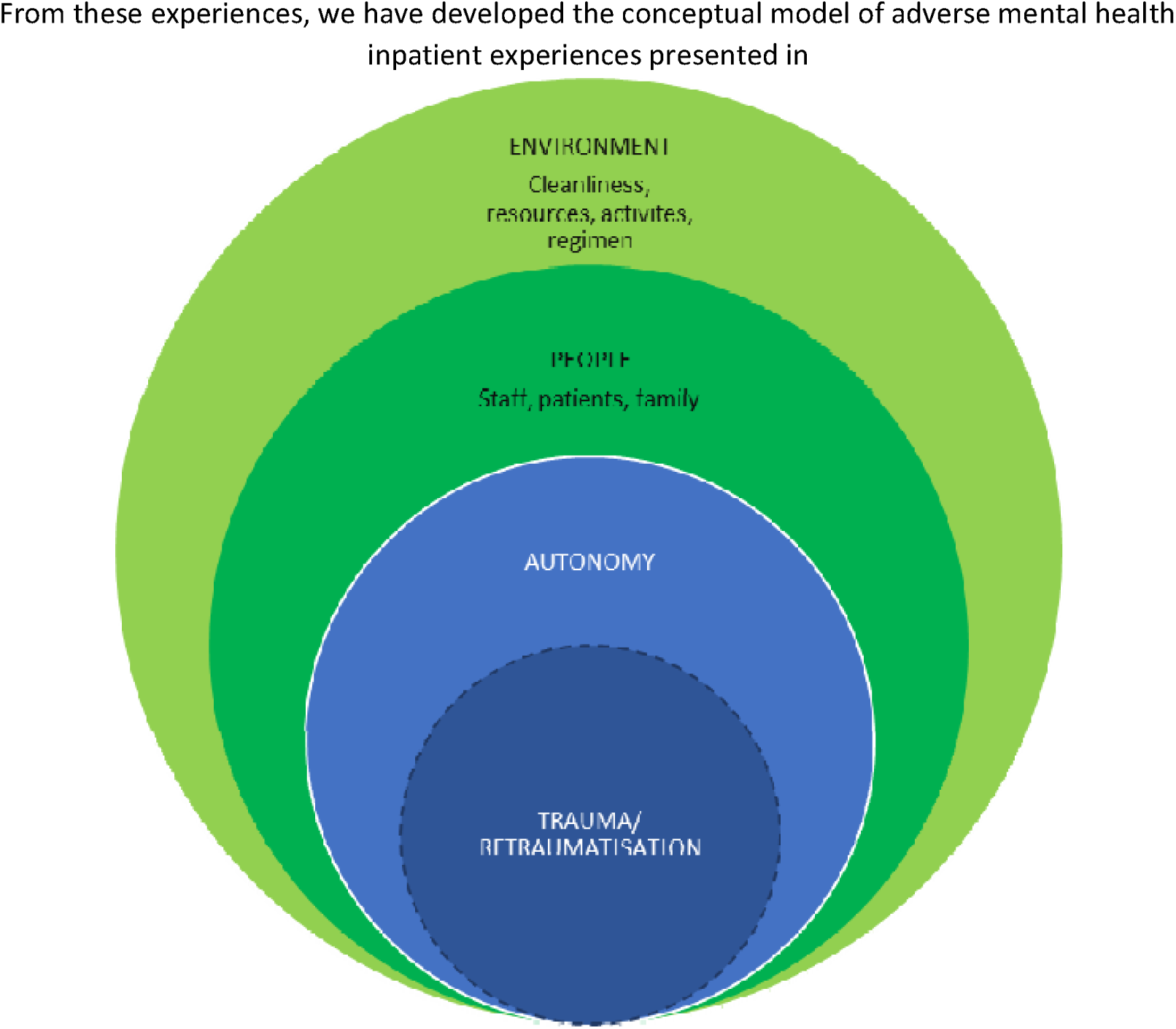
This reflects the themes evident in the literature and their putative relationship. The model comprises the ecosystem which represents the context within which adverse patient experiences occur. Environmental and person-related aspects may per se be related to adverse experiences or may exacerbate other experiences. Working across the ecosystem are the systems that operationalise mental health care and treatment including condoned practices such as seclusion and restraint, legislative aspects (detention, continued detentions) and formal processes (ward rounds). Given that these aspects are largely conducted by and within the ecosystem they are inherently connected. In addition is the changing context provided by transitions into, within and out of services. At the centre of the adverse patient experience, existing in multiple negatively perceived contexts is the individual patient who experiences loss of autonomy, power imbalances, trauma and retraumatisation.

**Figure 2.**
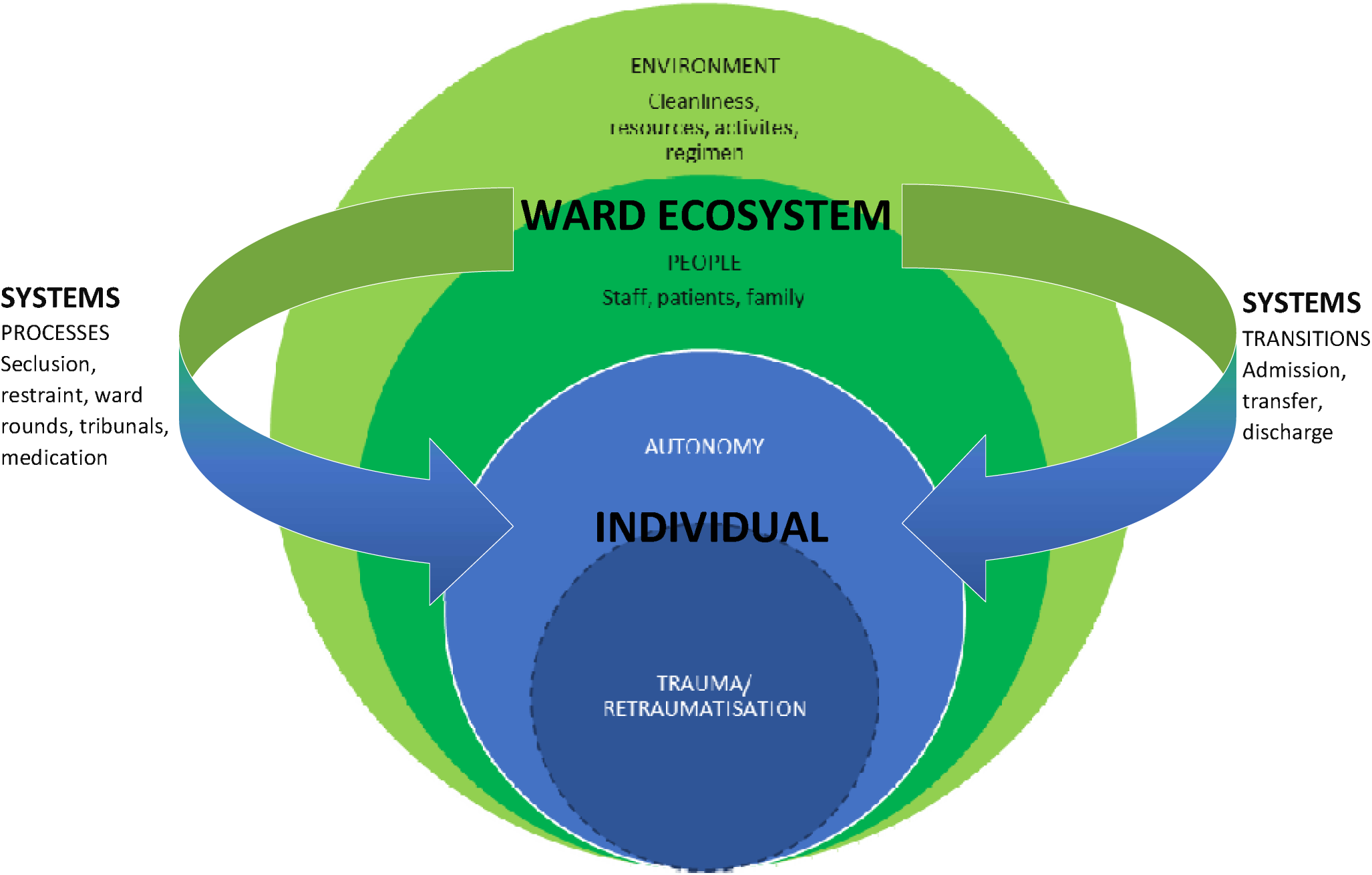
Conceptual framework of adverse mental health inpatient experiences.

This review has focused deliberately on negative experiences to synthesise a comprehensive overview and understanding of adverse experiences. In so doing it has highlighted both the role of the totality of the inpatient experience in adverse experiences and a broader range of singular experiences that can be perceived to be adverse by patients. This should not be taken to imply that patients do not have neutral or positive experiences. Neither should it be taken to imply that there can be no protective factors that might guard against adverse experiences and their impacts: such as, for example, ameliorating interventions to improve the environment, increase the capacity of people to provide a better experience, reduce coercive practices and provide better information. In addition, though not primarily, there may be individual factors for the patient that heighten the risk of adversity and its consequent impacts.

We propose that adversity develops across the domains (personal, system and external) rather than being distinct and separate. For example, restraint, which is within system processes, can cause trauma. It is worth noting, however, that not all patients who experience restraint also experience restraint-related trauma. This conceptual model can therefore be explored in relation to theories of resilience, which have their roots in the study of adversity.

Resilience research focuses on the processes that enable people to achieve better-than-expected outcomes in the face of adversity.^122^ Early research focused on individual factors, such as hardiness and self-efficacy, but has been criticised for aligning with neoliberalism and decentralisation of responsibility while disregarding social factors.^123^ This decentralisation of responsibility can be seen as a way for those in power to disregard adverse social systems and dynamics, such as poverty, racism, lack of access to resources and poor-quality education. Bottrell^124^^p335^ raises the notion of resistance within resilience theory, asking ‘“How much adversity should resilient individuals endure before social arrangements rather than individuals are targeted for intervention?’. van Breder^122^^p9^ develops this argument stating that ‘In the context of structural inequality, resistance to adversity is more appropriate than resilience’.

Yet, within mental health services, there exists an underlying expectation of resilience. This review has shown, that for some people at least, their mental health improves despite, rather than due to, their engagement with inpatient services. The current system often places excessive emphasis on the individual’s capacity for resilience, without taking into account the broader social context they find themselves in. How can someone expect to recover, or at least improve to the point of discharge, when they are surrounded by an ecosystem, and the associated processes and transitions, that create adversity?

It is noteworthy that in a phenomenological study of resilience in the lives of people who have experienced mental illness, acceptance of self, others and situation was described as integral to being resilient.^125^ Perhaps accepting that services create adversity is what allows people to recover enough to be discharged. Whether this hinges on acceptance or other mechanisms, it is plausible that mitigating the adversities individuals face could markedly enhance their potential for recovery. One resilience theory posits that resilience is the interplay between the individual, adversity and positive outcomes;^126^ could reducing inpatient adversities increase resilience? It is only really possible to answer this question once inpatient adversities are understood. The conceptual model presented here serves as a foundation for this understanding.

We have highlighted that inpatient adversity is more wide-ranging than either restrictive interventions alone, or even coercion more broadly, and the international span of available research suggests that it is a phenomenon that is not restricted to one country or even continent. When exploring how to best utilise this understanding of adversity in mental health inpatient settings, it is helpful to consider how adversity has been linked to health and wellbeing in other areas. Since Felitti et al^127^ developed the ACEs questionnaire in the late 1990s, understanding of the links between adversity in childhood and long-term outcomes have proliferated.^128^ Indeed there are a myriad of dimensional models specifying mechanistic pathways by which various dimensions of adversity are linked to health and wellbeing outcomes later in life.^129^ Measuring inpatient adversity might give a similar opportunity to specify mechanistic pathways that link to mental health outcomes. This has service-wide implications; increasing understanding of the impact of inpatient adversity will enable services to focus resources on the areas that will have the greatest impact both in the short-and long-term.

Moreover, differences in rates suggest that there is ethnicity-based inequity in hospital admissions for mental disorders. Hospitalisation for indigenous people in Australia is almost double that of other Australians^130^ while compulsory hospital admission is disproportionately experienced by Black patients.^131^ And once in services, there is further evidence of ethnicity-based inequity. In the UK, black patients are more likely to be restrained in the prone position than their white counterparts^131^, and similarly, in the USA, black patients are more likely to be physically and chemically restrained.^132^ It is possible, therefore, that inequity extends to all inpatient adversity, but without the means to measure it there is no way of determining the fact or extent of this claim. This review provides a framework from which to begin to understand and measure inpatient adversity.

### Strengths and limitations

This was an extensive review of 111 studies from 25 countries, involving > 4,000 individuals. This is, to the best of our knowledge, the first review to attempt to synthesise the extant literature on all adverse mental health inpatient experiences, rather than on coercion-related experience. While this is a strength, we acknowledge that in developing the search terms we focused on what is already known meaning that we missed search terms that may have produced studies that elicited greater depth, particularly in some areas, e.g. tribunals and ward rounds. However, we did not seek to quantify the experiences but instead intended to identify the spectrum of adverse experiences.

A common criticism of secondary research is that by its nature, it is reliant on available evidence, whose conduct and content may include biases that we could not address. Few studies, other than those on restrictive interventions, set out to capture people’s adverse experiences so it is possible that not all adverse experiences have been captured by this review. This is further compounded by the lack of patient voice in the studies; the findings from this review are based on participant quotes, however few, if any of the included studies, included patient co-researchers. This means that studies, and by extension their findings, may be influenced by researcher (and publication) biases. It is of note, for example, that racism was only identified in five studies and so could easily have been missed as a theme; we believe that the scant space given to racism reflects the research, not people’s experiences.

### Conclusions

This review has shown that, on a global scale, adversity reaches far beyond the harm caused by restrictive interventions. The conceptual framework demonstrates the interplay between the individual and the ward ecosystem and how they are affected by processes and transitions. This has important implications for service design and delivery. Improved patient experience is associated with improved patient outcomes, and addressing these negative experiences could significantly impact patient care. Mental health inpatient settings should strive to create an environment that is supportive, respectful, and safe for patients, considering the conceptual framework of adversity developed from this review. Research is needed to enable valid measurement of all inpatient adversity; it is only by understanding a problem that solutions can be found. This review clearly shows that some services are failing some patients and the similarities of experiences across borders suggest that these are not isolated incidents. We need to understand the interplay between inpatient adversities as well as the short-and long-term outcomes to support the development of services that work for the people in them.

## Data Availability

All data produced in the present study are available upon reasonable request to the authors

## Acknowledgements

We would like to thank the people who participated in the public and patient involvement activities.

## Author contributions

Nutmeg Hallett and Geoffrey L. Dickens have made substantial contributions to the conception or design of the work.

Nutmeg Hallett, Rachel Dickinson and Emachi Eneje have all made substantial contributions to the acquisition, analysis or interpretation of the data.

All authors have been involved in drafting the work or revising it critically for important intellectual content, have had final approval of the version to be published and agree to be accountable for all aspects of the work in ensuring that questions related to accuracy or integrity of any part of the work are appropriately investigated and resolved.

## Data availability

Data availability is not applicable to this article as no new data were created or analysed in this study.

