## Supplementary data for "Adverse mental health inpatient experiences: Qualitative systematic review of international literature and development of a conceptual framework"

Supplementary Table 1. Search terms

| **Database** | **Search terms*** |
| --- | --- |
| CINAHL Plus | (inpatients [SH] OR inpatients OR patients [SH] OR service user) AND (psychiatric commitment [SH] OR hospitals, psychiatric [SH] OR psychiatric units [SH] OR psychiatric OR psychiatry [SH] OR psychiatry OR mental disorders [SH]) AND patient seclusion [SH] OR seclusion OR restraint, chemical [SH] OR restraint, physical [SH] OR restraint OR tranquilli?ation OR sedation [SH] OR sedation OR involuntary treatment [SH] OR involuntary commitment [SH] OR involuntary OR police OR taser OR fear [SH] OR fear OR negative OR harm OR harmful OR coercive OR coercion) AND (patient satisfaction [SH] OR patient satisfaction OR satisfy* OR experien*) |
| Google Scholar | (Inpatients OR Patients) AND (psychiatric hospital OR psychiatry OR mental disorders) AND (seclusion OR restraint OR coercion OR forced medication OR police OR involuntary OR fear OR harm) AND (satisfaction or experiences) |
| Medline OVID | (inpatients OR inpatients [SH] OR service user OR patients [SH] OR commitment of mentally ill [SH]) AND (psychiatry [SH] OR psychiatry OR psychiatric OR hospitals, psychiatric [SH] OR mental disorders [SH]) AND (seclusion OR restraint, physical [SH] OR restraint OR tranquilli?ation OR sedation OR involuntary treatment [SH] OR involuntary commitment [SH] OR involuntary OR police OR taser OR fear OR negative OR harmful OR coercion OR coercive OR forced medication OR harm OR harmful) AND (patient satisfaction [SH] OR patient satisfaction OR satis* OR experien*) |
| PsycINFO | (psychiatric patients [SH] OR hospitalized patients [SH] OR psychiatric hospitalization [SH] OR inpatients OR service user OR patients [SH] OR commitment, psychiatric [SH]) AND (psychiatric hospitals [SH] OR psychiatry [SH] OR psychiatry OR mental disorders [SH]) AND (patient seclusion [SH] OR seclusion OR physical restraint [SH] OR restraint OR tranquilli?ation OR sedation OR involuntary treatment [SH] OR forced medication OR police personnel [SH] OR police OR law enforcement [SH] OR taser OR fear [SH] OR fear OR negative OR harm OR harmful OR coercive OR coercion) AND patient satisfaction OR client satisfaction [SH] OR satis* OR experien*) |

* Search terms are keywords unless labelled [SH] for subject heading

Supplementary Table 2. Study characteristics

| **Citation**  **Country** | **Purpose of study** | **Study design**  **Data collection,**  **data analysis** | **Time of data collection** | **Setting** | **Participants** | **Authors’ themes (in bold)/results with illustrative quotes (in italics) as relevant to the review** |
| --- | --- | --- | --- | --- | --- | --- |
| Ådnanes et al 2018  Romania, Slovenia, Finland, Italy, Austria, Norway | To describe and explore the psychiatric patient experience of rehospitalisation | Eight semi structured focus groups | In the community following at least one admission | Held in the community, based in large cities in each participating country | 55 participants, all in receipt of mental health services with one or more psychiatric hospitalisations | **Re-hospitalisation as less traumatising than the first hospitalisation**  *“For me, my first admission…it was… It was a trauma, a very big one. I knew what I wanted, I was fit to plead, but it was something ‘induced’ by someone else.”*  *“I must say that now - the second time around was better.”*  **Re-hospitalisation as inevitable and by default but without progress**  *“Well, people come in again and again and again because psychiatry “chronicises” instead of helping people regain their health.”*  **Re-hospitalisation as part of a recovery process?**  *“You are by yourself too much [in the hospital]. Maybe some psychotherapy would be good. That would be necessary to implement. I think there is not enough of it.”* |
| Alexander 2006  UK | To explore patient feelings about ward nursing regimes and involvement in rule construction | Semi-structured interviews | During admission | Two acute psychiatric wards | 30 patients admitted to psychiatric acute wards | **Coercion**  *I don’t know where I stand here psychologically it is extremely in the back of my mind its, its it permeates my being this unsteady feeling of terror, because I never know what to expect, or what’s going to happen. They give you the impression that everything’s so fine and easy going, but then they suddenly do such strange, and take such strange measures*  **Distress**  *This one particular nurse she says to me, do you understand why I am tell-ing you this? I said no I don’t, you are picking on me, you should pick on other patients for a change.*  **Confinement**  *I dislike the locked door; it stands symbolically to me that it’s just possible you may never get out. Because in that way if they don’t behave themselves, they won’t get out.*  **Acceptance**  *To get into bed dirty it’s not me it’s not my style. [Inter-viewer: How do you feel when you are refused permission to do something?] Just accepting that you are mentally ill anyway and you get told you can’t do that.*  **Humiliation**  *I was humiliated yesterday by one of the staff, even though I had been pestering her before, she might have been fed up with me, and she told me to get out!*  **Anger**  *You can give your opinion, but then if you get frustrated and angry because their opinion is the only opinion that counts when they are giving out medication. If you show any anger then they naturally say, well all right you’re not well, it’s this, it’s that, and they try and tell you what you are thinking and feelings* |
| Allikmets et al 2020  UK | To describe the patient experience of seclusion | Phenomenology Structured 1-1 interviews | During admission | Psychiatric intensive care unit | 10 male patients aged 18-25, on PICU, detained under section 2 or 3 MHA | **Physical aggression against patients**  Staff were *too painful* and *too rough* during incidents leading to seclusion  *They never talked to you. When you have people come into the cell with a gang of people, I’m defensive straight away. I was erratic but I believe a conversation should have happened. Felt people ganged up on me and tried to force me to do something. I felt threatened by forced medication.*  **Lack of social and psychological support**  *Scared they were going to inject me more. I never get told how long I will be there, and I can’t phone my family.*  **Need for improving/replacing the practice of seclusion**  *I think the staff bully you … If in supervised confinement, you should be allowed newspapers/books or a bible. It’s boring, you end up going mad.* |
| Askew et al 2019  UK | To describe and explore the forensic patient experience of seclusion | Interpretative phenomenological analysis  Semi-structured interviews | During admission, at least 28+ days following episode of seclusion | Forensic inpatient psychiatric unit | Seven male forensic patients across 3 medium secure wards | **Intense fear**  *Every time they open the door, they kinda like all in gloves and there was about 12 of them, I thought, what the fuck’s going on here, that’s why I was getting you know like ideas in my head thinking they’re gonna fuckin’ kill me*  **Not getting the care I needed**  *Left in a seclusion room for a week without my clothes. I shit up the walls*  **I am being abused**  *Felt like I was being like, visually abused or something. It didn't feel, feeling right at all*  **Power struggle**  *In a place where all the control is taken off you, yeah, you’ve suddenly got a bit of control. You’re in an environment where you don’t have any control, everybody’s trying to grab that little bit of control* |
| Askola et al 2016  Finland | To explore forensic patients’ experience of their treatment and working through the offence | Semi-structured interviews, narrative analysis | During admission, however two patients had been discharged by National Institute for Health and Welfare under supervision and were residing in psychiatric rehabilitation units | Finnish forensic psychiatric hospital | Eight patients (seven males, one female) all aged between 30-50.  All had been in a psychiatric forensic hospital, though two had moved to a less restrictive rehabilitation unit | **Involuntary admission**  *Then the forensic psychiatric examination began and it was pretty tough when they took away my medication and I had an awful lot of medications, I was in poor condition, I was a polydrug user, and I had withdrawal symptoms on top of it all.*  **Life as a forensic psychiatric patient in an institution**  *I was in mechanical restraints and pulled the straps off. I had straps on my wrists and I broke them, I had incredible strength. Then they put other straps on me, and they broke, too, they said that if I didn’t calm down they would put me all in straps, which means that they would put them all round my body, like winding sheets. Then they put this sheet on my head so you don’t see what’s going on. When I was secluded they never came to say come here then, let’s go and play.*  **Trauma, resilience and recovery**  *Life gives hope. I have my own plans but I keep them to myself. But I can say that they are all within the law. We surely all have our own goals where we want to get, but you don’t need think about them all the time.* |
| Barnicot et al 2016  UK | Staff and patient experiences of decision making regarding continuous observations in psychiatric hospitals | Thematic analysis of semi-structured interviews | During admission | Two acute psychiatric hospital wards, London | 28 inpatients who had been on continuous observations within the past year | **The conflict between privacy and safety**  *You lose your space and it feels like you’re being invaded and they’re in control of you.*  **A damaging intervention versus a short-term solution within a positive risk-taking framework**  *I hate being stared at. It makes me agitated.....It made me do it [self-harm] more.... because of the amount of pressure they put on me.*  **Decisions made without the patient versus a collaborative and individualised approach**  *I knew that I wasn’t a danger to myself ... I was in a safe environment.... I didn’t need to be watched... I didn’t see the sense in it.*  **A stressed and fractured workforce versus a team approach**  *Slap-dash approach, leaving me alone, wouldn’t bother to do the one-to-one half the time...... It really upset me because I felt really judged... the conflict between nurses made me feel worse.* |
| Bonner et al 2002  UK | To explore staff and patient experience of physical restraint | Pilot study, semi structured interviews | As soon as possible following incidents | Acute psychiatric ward | Six patients, demographics unknown | **Antecedents**  *I got angry because they wouldn’t listen to what I was trying to tell them. Telling them that I needed help, wanted to hurt myself . . . it was horrible, I never want it to happen again*  **In the midst of conflict**  *Disgusted that a male nurse was present. . . it was bad enough having the injection without the embarrassment of having a male nurse present. Only female nurses should be present when restraint or injections are given to female patients*  **The aftermath**  *I don’t know why they didn’t sit down and talk to me. I’d been in a mute state. I thought that they’d try to come and speak to me. After [the incident] a student nurse came and spoke to me. It was the first conversation I had in days. I found that helpful. The ward staff involved were unapproachable*  **Other issues**  *My biggest fear before coming into hospital is being restrained. It puts me off seeking helped because I’m frightened. I then become ill and end up being admitted and restrained anyway.*  *They [the agency staff] are only in it for the money. They sit and watch telly, play pool and basically ignore the patients* |
| Bradbury et al 2016  Australia | To understand the lived experience of involuntary transport under MHA from the perspectives of consumers, carers, mental health nurses, police officers, and ambulatory paramedics | Cross sectional qualitative review, using semi structured interviews | In the community following discharge | At participants convenience – university campus, hospital, and participants place of home and work | Six mental health consumers with experience of involuntary transportation under MHA | **Humiliated by usually harsh treatment**  *There’s shame and criminalization and acute embarrassment and mortification in being picked up by the police for being ill (sic)...They were very gentle. You know, you are (name of person), you must come with us. Open the paddy wagon, you know, hand under the elbow, in, but it’s still a paddy wagon. It’s still two coppers. It’s not anybody with clinical skill. It’s not anybody necessarily with much kindness, because he’s a law enforcement person* |
| Buizza et al 2007  Italy | To identify and understand the stigma from the perspective of people with schizophrenia and their relatives in local context | Focus groups | During active treatment, however not all patient participants were inpatient at time of focus groups | Psychiatric rehabilitation unit | 26 patients across six focus groups. All patients were in treatment at a psychiatric rehabilitation unit, 75% inpatient and 25% outpatient | **Quality of mental health services**  *They are going to open a sheltered apartment in street G. I ask myself: Does it have to be in the industrial area at the outskirts of this town? There aren't any services, facilities or stores, there!* |
| Burn et al 2019  UK | How do patients and clinicians experience the OPeNS intervention session and how many patients are able to participate in the OPeNS intervention within 1 week of admission | Mixed methodology, semi structured interviews, Clients Assessment of Treatment Scale and clinician collected data | Within the first week of involuntary admission under MHA | Acute psychiatric ward | 14 patients, all detained under MHA on an acute psychiatric ward | **Clashing with usual practices and priorities**  *The staff are too busy. There’s about five patients per staff. You should have seen yesterday, there’s only two staff to ten patients.*  *It’s not the environment it’s just the attitude to work.* |
| Chambers et al 2014  UK | To explore service user experience of detained care, reflecting on dignity and respect and coercive interventions | Semi structured interviews, thematic analysis | During admission. All participants had been detained for between two weeks and two months | Three mental health hospitals – an acute ward, a PICU and a forensic ward were included | 19 patients detained under MHA on acute, PICU or forensic wards | **Heard by staff members**  *But overall, they’re together, kind of, and they do things against you. Most of them don’t even care, you know? They don’t seem to have much love or respect for you.*  **Involvement in decision making regarding their care**  *Definitely restricted (with regards to medication treatment), but coerced into taking medication. Cos nothing changes, when you ask for something to change, like medication, nothing changes until they’re ready to change you.*  **Information about their treatment plans**  *No-one was explaining to me what was happening. Uh, it was just given to me (medication); nobody did sit and discuss it with me.*  **Coercive interventions and alternatives**  *Pumped with more medication that they don’t need*  *When they dragged me forward, one of them had to drag me by my trousers… And they pulled them down, and left them. That’s out of order. It depends how it happens, some of them can be really embarrassing. Like me, I was getting dragged along in my underpants*  *Well, the scariest time was the first time, I was just petrified. And then the team came in and they held me down and um, they gave me an injection, and I was very scared*  *Some of them can be cruel, you know? They pull their clothes around their neck and they’re almost strangling them, and they hold their hand behind their back and it’s cruel and they’re squeezing their arms. Some of them can be very cruel…. Some of them take unfair advantage*  **Physical setting/environment and daily activities**  *There’s less, uh, restrictions in prison, you know? Like, we feel like we’re in prison but they call it a hospital* |
| Cheetham et al 2017  UK | To explore how patients discuss their inpatient experiences and how it relates to their social position on the unit | Foucauldian discourse analysis, focus groups and semi structured interviews | Minimum of six months+ following discharge from inpatient care | Community, though unclear where | Nine service users participated.  All nine attended the focus group.  Two of these nine were also interviewed | **Medical-technical-legal**  *I think with the staff there, it’s very plastic. It’s very false. There is no relationship…there’s a glass wall between us.*  **Ordinary humane relating**  *[staff] are treating the patients like children as well, you know, like a teacher would be quite rude to students in terms of, sort of shouting at them and telling them what to do and then expect the students to be polite back. It sort of, seems to be like either a parent and a child or a teacher-student relationship. They’re not treating patients like other adults that deserve a certain amount of respect.*  *[Nurses]They’re just ticking boxes, doing paperwork. They’ve got no time for you [...] [Healthcare Assistants] they’re the ones that are more human ‘cos they’ve not been programmed yet****.*** |
| Chien et al 2005  Hong Kong | To explore patient perception of physical restraint in psychiatric inpatient care | Qualitative exploratory study, semi structured interviews | Less than two days following first instance of restraint | Acute psychiatric inpatient ward | 30 patients, 18 male & 12 female from acute psychiatric wards, who experienced their first restraint less than two days prior to interview | **Negative and non-therapeutic impacts of restraint**  **1. Lack of concern and empathy**  *At times my existence was ignored... no matter what I said to the staff passing by, they did not stop, or respond to my requests. I felt that I did not exist over there.*  **2. Powerlessness and uncertainty**  *I was afraid that someone might hurt me suddenly while I was being restrained. Being restrained could be terrible if you did not know for sure what would happen next... especially at night, or over a long period. I was scared... nobody seemed willing to help me calm this fear and I was afraid the staff were never going to take the restraint off.* |
| Chorlton et al 2015  UK | To explore how illicit substance users experience relationships with staff during psychiatric inpatient admissions | Interpretative phenomenological analysis, semi structured interviews | During admission | Eight psychiatric inpatient wards | Self-reported illicit substance users currently detained on an acute psychiatric ward. Ten patients, five male and five female participated | **Weighing up the risks of relationships**  *There are some you can’t [approach], because you don’t feel […] that you’re going to get the reaction you want.*  **Relationships intertwined with power and control**  *It’s just like being in gaol. I couldn’t go out.*  *I was swearing that much that I ended up getting restrained, and then that was it, I didn’t speak to her for 2 months’.*  **Seeking compassionate care**  *Some of them just talk to you like you’re [expletive]. […] Like they don’t care’.*  *I’ve been thinking ‘oh no I’m getting discharged on Tuesday and I’ve got nothing in place’ […] And then I got frustrated and upset about it and I went out and had a drink’.* |
| Cutcliffe et al 2012  Canada | To explore the increased risk for suicide following discharge from inpatient psychiatric admission | Mixed methods phenomenological study, using semi structured interviews | At one month following discharge from inpatient services | In the community, place chosen by each participant | 20 participants (10 male and 10 female). All participants had a history of suicidal ideation/behaviour and had been discharged from inpatient care one month previously | **Existential angst at the prospect of discharge**  *I was like, Oh my God, I’m going back to the same thing where nothing has really changed.*  *I like it here but I can’t stay here forever.*  **Preparedness**  *Yeah, the psychiatrist I had in there wasn’t listening to me anyways, and she had her own protocols and she said I was being discharged and it’s like all the time I was there treatment-wise, you know, I was telling her that I was still not feeling well, and you know you don’t know say that with suicide.* |
| Cutting and Henderson 2002  UK | To explore women’s experiences of inpatient psychiatric services | Grounded theory, semi structured interviews and focus groups | Unclear. Some were held in the community after discharge. Interviews took place at hospital - it is not clear whether they were inpatient or outpatient participants.  Hospital was mixed gender ward. | In the community, day/resource centres and at the hospital site | Eight interviews took place.  Two focus groups took place, unclear number of participants.  No demographic data of participants. | **A different world**  *It’s very punitive and very stressful. I keep my mouth shut*  *The atmosphere was so depressing, there didn’t appear to be anything to do, the physical environment was very oppressive.*  *It’s a pigsty. It’s filthy, it’s hideous . . . it’s just horrible... put people in that environment and they will act accordingly.*  **A woman in a man’s world**  *I don’t think that there was any loving care there...the people that are there are called caring assistants, I didn’t feel they were caring at all.*  *There were very few staff around, it felt very unsafe, I thought I would be in more danger going there than I would be staying out.*  **A pill for every ill**  *She discharged me that day even though there were housing issues that hadn’t been sorted out. I asked if I could have a couple of days leeway but she said no, I was to be discharged there and then and just led on to the homeless person unit which I had to go to. I was put into the most grotty bed and breakfast, I was given a fortnight’s medication... Things looked really bleak and that night I took an overdose of all my medication.*  **Will you hear us**  *I felt like killing myself last night...and I came in really frightened and you know just trying to speak to somebody...it would be easier to sit down and talk to one person than having snatched bits of conversation with people who then have to leave you half way through to run off and go and see to someone else just having a bit of quality time I think.* |
| De Pau et al 2020  Belgium | To explore the experience of receiving care when labelled as not criminally responsible, particularly their care trajectories | Interpretative, semi structured interviews | During admission. One participant and been in hospital for less than one year. The longest interment was for 27 years. | Low, medium and high secure hospitals across Belgium | Five female and 18 male forensic patients from low, medium and high secure forensic inpatient wards.  Participants had a range of diagnoses, a range of length of interment and index offences | **Decision making process**  *It is a small group of five people who were present during the hearing and who made a decision, between them, without involving me.*  **Transition moments**  *They never gave me the information. They opened the door, they let me go. I took the train and went home.*  **Structural factors influencing care trajectories**  *And then you don't know what to do anymore. I stayed two more years in this setting because he [the social worker] didn't send the file and I couldn't appeal it because of the transition to the new law.*  **Relation to time**  *I know people who stay here for eighteen or nineteen years. Please, don't do that to me.* |
| Desplenter et al 2013  Belgium | To explore practices regarding the provision of information about antidepressants to patients with depression admitted to psychiatric hospital | Qualitative semi structured interviews | During admission, patients nearing the point of discharge | Eight psychiatric hospitals across Flanders | 17 patients (14 female and three male). All were admitted with mood disorders and had been identified as being close to discharge | **Medication information on demand of the patient**  *Yes uh, but not so often in fact. It happens that people ask for information, euh … but there are also people with many questions who finally will not dare to ask them.* |
| Duggins and Shaw 2006  UK | To examine the concept of patient satisfaction in people with schizophrenia in the context of a recent inpatient admission | Cognitive mapping, semi structured interviews | Within one year of discharge from acute inpatient care | In the community, often participants own homes | Ten participants (six male and four female). Seven had been detained under MHA, three had been informal. Wide range of ages, ethnicities and number of previous admissions | **External factors**  *9 times out of 10 whenever I’ve been admitted or admitted myself there are people that are physically violent, threatening towards other people.*  *It would have been a lot, a lot easier for me if they had of given some kind of induction. Because when I got there basically it’s just they showed me where to smoke and where to sit and watch TV and showed me my room and that was it. They went off and did their thing and I was left to just wander about basically.*  *People run your life for you. You don’t run your own life.*  **Internal factors**  *From a Christian point of view, I try to look at it as something spiritual or a spiritual experience. I could have dealt with it from that angle, but because I was pulled out of that sort of environment, and put into another, one where the emphasis was on the treatment, and like the medication.* |
| Efkemann et al 2019  Germany | To explore the experience of patients and staff experiences to open and locked wards, relating this to ward atmosphere and patient safety | Mixed methods, quantitative questionnaire and qualitative semi structured interviews | During admission | A facultative locked hospital in Germany | 15 patients on a facultative locked acute ward.  No demographics provided | **Patients’ cohesion**  *Well, simply that patients, loosely speaking, were threatening or bullying one another or whatever.*  **Experienced safety**  *You build up frustration not only as a reasonably healthy but also as an ill person. You can’t get out. The door is locked. And then frustration comes and … trash, littering, or demolishing walls or I don’t know. Frustration bubbles up because the door is locked.*  **Patient satisfaction**  No quotes presented. |
| El-Badri and Mellsop 2008  New Zealand | To explore the perceptions and experience of patients and staff on the use of seclusion in psychiatric services | Mixed methods questionnaires | In the community, unclear at what point following discharge | Psychiatric outpatient clinics | 111 patients completed the questionnaire.  Age range 18-65 | **Use of seclusion**  *Forced seclusion is mainly a way for those in authority to gain control over what they deem to be unsatisfactory behaviour in patients.*  **Emotional impact**  *Terrified not being told why I had been locked in this dark room on a mattress on the floor. I felt like a caged animal and wondered why my family could not come in and visit with me’.*  *I hate the conditions of seclusion e.g. mattress, no toilet ... this is inhumane and obsolete treatment.* |
| Ezeobele et al 2014  USA | To explore and describe the psychiatric patients’ lived seclusion experience | Phenomenological framework, semi structured interviews | On the ward, three days following discontinuation of seclusion | Acute psychiatric inpatient ward | 20 participants detained on an acute psychiatric ward who had been secluded three days prior to interview.  12 male, eight female participants.  Age range 19-53 | **Alone in the world**  *I cannot get things that I want to do or get out, nobody was listening, the doors are shut behind you, and you are there alone.*  *I felt violated . . . I felt everything had been stripped from me . . . I felt ashamed because I wanted to cooperate with the staff, you know . . .*  **Staff exert power and control**  *The nurse told me to take my medicines…the nurse did not explain the situation to me…rather…uh…the nurse called four big guys and they held me…the nurse refused to listen to me. I was afraid and powerless. I did not know what they were going to do to me. They outnumbered me.*  **Resentment towards staff**  *The nurses hollered at me…spoke to me in a derogatory tone and made jokes. The nurse should have listened to me.* |
| Faschingbauer et al 2013  USA | To explore the lived experience of inpatient psychiatric patients who are placed in seclusion | Phenomenological approach, unstructured interviews | Between one-seven days of discontinuation of seclusion | Psychiatric inpatient ward | 12 participants (six male and six female)  Age range 18-50  Range of ethnicities and diagnoses | **Patient hope for respect and open communication**  *I know that I don’t like to be told what to do. I like to be asked if I would do something, rather than told to do it. That approach to me would have been able to avoid seclusion completely.*  **Patient emotional response to the seclusion process**  *They would not do anything for me, they just kind of basically were laughing. That almost set me off again, because, you know, these are your nurses, they are supposed to be taking care of you, and you don’t feel like you are being taken care of when someone is making fun or laughing at your situation.*  **Patient Insight into Behavior and the Importance of Positive Coping Skills**  *But, before they put me in there, I was just trying to blow off steam. In my workbooks, it said you could punch a pillow if you feel frustrated. That is what I was doing, and they had a problem with that, so then they put me in there.* |
| Fenton et al 2014  UK | To explore the experience of hospitalisation for early psychosis | Interpretative phenomenological analysis, semi structured interviews | In the community following discharge | In participants own homes | Six participants (five male and one female), aged 18-33.  All had been admitted at least once.  All were under Early Intervention Services | **Confusion and uncertainty**  *I just thought, ‘I’m gonna die.” I was in the back of the van going somewhere and I didn’t know where I was going - that was when I became quite panicky and obviously I got took to the [unit] which I didn’t know was the [unit]. I hadn’t got a clue, all I knew was that the police was the highest authority at the time and they were taking me somewhere else.*  **Feeling chaotic and unsafe**  *I would classify my whole experience in there as quite horrific […] it was just horrible being in there.*  **Maintaining identity**  *The people there they were so, so heavily you know in psychosis it was very disturbing to be around them and feel like I’m in the same place as these people, and you feel like you are being classified in the same way as them and it scares you a bit as you almost feel like somehow being around them is going to make you worse.* |
| Fixsen 2021  UK | To interpret the researchers’ own experiences as a patient in a psychiatric ward during the COVID-19 lockdown | Interpretative autoethnography, emotional recall | Following discharge | Acute psychiatric ward | Author | **No themes**  *I have come to accept such things [coercion] with an inevitability that is uncharacteristic, but when the alternative of being forcibly injected with the prescribed drugs was presented to me, I caved in.* |
| Fletcher et al 2019  Australia | To describe the impact of Safewards on patient experience in inpatient mental health services | Mixed methods, postintervention survey | During admission | 10 inpatient mental health wards | 72 inpatients completed the survey (29 male and 31 female), age range 18-78 | **Patronising language and intention**  *I don’t think it’s respectful to treat people as a child.* |
| Fortune et al 2010  UK | To describe the experiences of staff and service users in personality disorder offenders’ forensic services | Semi structured interviews and focus groups | During admission | Three inpatient medium secure forensic mental health services | 30 service users across the three services.  All had primary diagnosis of personality disorder.  Age range 22-56  Length of treatment range 1-23 months | **The experience of receiving treatment**  *People are at us all the time about how I feel, “How do you feel about this?”, “How do you feel about that?”. You know, have a conversation, “What do you feel?” – bombarded by “how you feel” questions, you know it’s just... I know why they are doing it, because the logic is not enough to stop us from getting in trouble again to have an emotional connection to what I have done in the past with my crimes and to have an emotional connection between what’s wrong and right makes a difference in whether or not I will reoffend again. But I know why they are doing it, but it doesn’t make it easy. Does my head in some days you know, really does my head in.*  **Delivering treatment**  *In a place like this you really need regular staff who are there day in day out, who can build up a rapport with the patients and who the patients feel comfortable in approaching to talk to about problems. Here, you’ve got a lot of staff who you might see every couple of months, like because they are not the nursing bank. And you’ve also got a lot of the bank staff who basically aren’t interested in doing anything other than the minimum amount possible and getting a pay packet at the end of the month... I feel that they are more committed to the pay packet than actually to you know, trying to help people.*  **Areas for improvement**  *You can cut it [the atmosphere] with a knife sometimes... between the staff and patients. And patients and patients. Because you have a mixture of prisoners and mental health patients. Prisoners are treated with no real self-importance. The mental health patients are treated like royalty...* |
| Fredriksen et al 2020  Norway | To explore the experience of patients with depressive psychosis in inpatient treatment, in relation to suicidal thoughts and behaviours | Semi structured interviews | Patients were enrolled in the study in first week of hospitalisation.  Interviews completed as approaching discharge | Psychiatric inpatient unit | Nine inpatients, (five female, four male)  Age range 19-55 | No relevant quotes. |
| Giacco et al 2018  UK | To identify barriers and facilitators to shared decision making with involuntary patients | Focus groups and semi structured interviews, thematic analysis | Some patients were interviewed whilst still detained in hospital.  Other focus groups and interviews took place in the community following discharge | Across different NHS trust sites, including community bases and hospital wards | Focus groups -  18 patient participants (10 female and eight male), within four months of their last involuntary admission.  1-1 interviews -  Six patient participants, all requiring an interpreter to allow participation | **Barriers to patient involvement in decisions**  *Just feel like they are being jailed and they don’t understand, you know. The staff described it as ‘detention’, for example, in a patient’s mind it’s like ‘I can’t leave’ or ‘it’s the same as being thrown into a cell’.*  **Facilitators of patient involvement in decisions**  *[The staff] some of them, they listen to what you want. Some of them they ignore what you are saying, absolutely … that makes it difficult.* |
| Gilburt et al 2008  UK | To explore the patient experience of admissions to acute psychiatric hospital | Participatory research approach, focus groups and semi structured interviews, thematic analysis | In the community, unclear time frame since discharge | Mental health resource centres | 19 participants (10 male, nine female), range of ethnicities. All had one or more admissions to inpatient services | **The role of communication**  *The staff work hard at trying to stay away from the clients was my opinion. Be in their office as much as they could.*  **Coercion**  *See the first time I ever went in there I think I was on a section actually and it felt horrible. It felt horrible because I was locked away for so many days and I couldn't go out and be free.*  *They wanted to tear me to pieces and I have arthritis of the shoulder to prove it.*  *Knowing that if I tried to leave I would just get sectioned. So it was a terrifying place, position to be in.*  *I was forced to take medication that was causing me a lot of discomfort.*  **Safety from self, safety from others**  *I was safe in hospital until somebody, some other patient tried to strangle me.*  *I felt frightened of the doctors, they were putting me on drugs that had terrible reactions. I felt frightened.*  **Treatment**  *I wasn't able to do anything, only take the tablets and be like a zombie all the time.*  **Cultural competency**  *You know you've got to be conscious of being black, you've got to be victimised for being black, and therefore we'll hurt you, intended to feel like this, because it's a kind of racism. ...And that's what I experienced in the psychiatric system.*  **Freedom**  *When I was first there I was distraught and what really distraughted me was when I weren't allowed to go outside and get a drink or anything like that.*  **Environment**  *It isn't nice, it's an absolute disgrace. There are no curtains, in the corridor or the smoking room. The windows are filthy; the furniture's filthy and burnt. It's an absolute dive. It's disgusting and I wouldn't put a pig there let alone a human being.* |
| Gilburt et al 2010  UK | To explore the subjective patient experience of traditional hospital services and residential alternatives | Qualitative, in-depth interviews | All participants were currently in one of the residential services described | Four crisis houses, a brief admission hospital ward and a traditional acute ward | 40 patients in residential alternative services who had previous experience of traditional hospital inpatient stays | **Opinions about services**  *It was a total and utter nightmare, I’ve never experienced anything so extreme with people running up and down corridors, screaming at 3 in the morning, I was terrified. I would never go there again. Hospital would make me worse.*  **Relationships**  *They were rude, their job was to, there was, although people were sick, right, they thought everybody was stupid. They’d talk down to us like we were lower than them.*  **Coercion**  *Yeah, forced, the medicine. They say if I don’t take the tablet they were going to inject me.*  **Freedom**  *You feel like a prisoner, that isn’t safe, you have to escape.*  **Paternalism**  *They literally treat you like babies in here. They feed you a two-hourly basis which is nice...then watch what you eat and they’ll make sure that you are looking after yourself. That’s how it should be.*  **Safety**  *Nurse X broke someone’s arm under restraint before I got here.*  *I felt the whole environment was very, very threatening . . . the nurses refusing to listen or understand*. |
| Hagen and Nixen 2011  Canada | To describe the experience of patients who self-identify as having recovered from psychosis | Phenomenological hermeneutic research design, narrative interview method | Average length of psychosis-free recovery was eight years. Substantial time since last admission | In the community, unclear | 18 participants, all female, age range 27-57  Average time free of psychosis – eight years | **Invalidated and unheard**  *They (the fellow patients) finally just told me, “Be careful what you speak out loud here, it’s not safe in here.”*  **Violence and violations**  *I found out pretty quickly that they could actually incarcerate you. The experience was one of being completely powerless, because you are powerless. Someone has signed you in, but you don’t have the power to get yourself out.*  *To be stripped naked and put in this stupid gown with your backside showing, put in restraints, and shot up the butt with a very serious dose of Stelazine. It’s a complete violation. It strips you literally of all your dignity. It is also really traumatic.* |
| Haglund et al 2003  Sweden | To describe patient experience of and nurse perception of forced medication | Explorative design, semi-structured interviews, content analysis | During admission, no specific timeframe following use of forced medication | Five locked psychiatric inpatient wards | 11 patients participated (eight female and three male)  All had psychotic disturbances at the time of forced medication  Age range 20-80 | **Resignation**  *You can’t do anything, it’s like being chained.*  **Violation of integrity**  *I am totally without legal rights.*  **Fear**  *I really believed I was going to die.*  **Retrospective approval [of forced medication]**  *Nobody has ever asked me why I have been crazy and mad when coming to the hospital, they have just injected Cisordinol, telling me that this is no family therapy.*  **Alternatives to forced medication**  *But I wish they had talked to me, I am not especially against medicines.* |
| Haglund and von Essen 2005  Sweden | To describe voluntary admitted patients’ perceptions of being cared for on a locked psychiatric ward and how they view this in relation to coercion | Explorative descriptive study, semi-structured interviews, content analysis | During admission | Seven psychiatric inpatient wards | 20 voluntarily admitted patients (10 male and 10 female), age range 19-87, current period of care ranged from 7-210 days. Patients had a range of diagnoses | **Confinement**  *I think it affects your self-confidence in the long term, as a person. I think that being ... being independent as a person, that is taken away from you. After all you are an adult who should manage on your own, and this makes it into something mental, which means that it is taken away from you and that can make you sensitive in this sort of situation.*  **Dependence**  *Having to ask to be able to go out, so that one of the staff can unlock the door. You can’t always find one of them. I feel as though I’m bothering them because I have to ask for help.*  **Feeling worse emotionally**  *Yeah, and there’s an uneasy feeling, a ‘disadvantage-feeling’.*  **Non-caring environment**  *The rattling of the keys, it’s a bit like being locked up in a prison’.*  **Concerns for visitors’ reactions**  *It makes you wonder what people you know might think about you when they come here and you are locked in.* |
| Haw et al 2011  UK | To explore forensic inpatients’ experiences of coercive treatments including physical restraint, seclusion and forced medication | Mixed methodology, written questionnaire, semi structured interviews, thematic analysis | During admission | Forensic psychiatric hospital, total 700 bed | 57 patients detained in adult forensic inpatient wards (27 males and 30 females), age range 19-52 | **Participants’ experiences of coercive treatments**  *It’s like a prison. It’s a small room with no ventilation, no window, no fan, no bed often, nothing.*  **Unpleasant physical environment**  *It was horrible in there, like rough sleeping for five days.*  **Unpleasant thoughts and emotions**  *It brings on intense feelings of shame, embarrassment and humiliation. It’s dehumanising.*  *When they strip you off even if you have a history of self harm they will strip you off. If you have had sexual abuse this is not very good.*  **Physical pain, injury and fear of death**  *They twist your wrists hard and dig their nails to put your face on the ground. They hold you down hard. You are screaming. They say calm down. It is harsh on us.*  **Control**  *You have no control. They have your hands, head down and you can’t see where you are going.*  **Loss of privileges**  *It makes you look bad. It makes it difficult to get out of here (hospital) and makes it difficult to be discharged.*  **Attitudes and experience of staff conducting coercive treatments**  *I end up feeling punished. They just want to keep me in there.*  *When they twist your arms and legs back it feels like they hate you.* |
| Hoekstra et al 2004  Netherlands | To explore the seclusion experience of chronic psychiatric patients | Grounded theory, semi structured interviews | Substantial time after admission (length of time not specified) | Patient homes and at treatment centres if chosen by participants | Seven participants, four females and three males, age range 29-41 | **Autonomy**  *You see, when I’m wearing a straitjacket there’s absolutely nothing I can do myself. I mean . . . when you’re restrained, you can’t do anything, you can’t even turn round.*  **Trust**  *I’m still having problems with enclosed spaces. I remember that at first . . . when I’d just got out of the . . . when it was about half a year ago, that I didn’t lock the loo and the shower at my parents’ place. That I just didn’t lock any door, so I could always get out. And that I couldn’t bear the sound of keys, a bunch of keys, you know, I’ll never forget the sound, this click of this heavy door and eh . . . well, it sticks in your mind.*  **Loneliness**  *A terrible feeling of loneliness. Especially these heavy doors and . . . they slam shut behind you and . . . I have never ever experienced such loneliness.* |
| Holmes et al 2014  Canada | To describe patients’ experience of seclusion | Hermeneutic phenomenological design, semi structured non directive interview, content analysis | During admission, within seven days of being secluded | Psychiatric inpatient hospital | Six patients, no demographics available | **Emotional impacts of the seclusion room experience**  *It was quiet, it was quiet. I wanted to get out of there because I was depressed to be alone, to be locked up. I was depressed from being alone, without people.*  *I have to get undressed in front of them. There are men and women. I’m totally naked and they put a johny shirt on me. They take everything off, everything. I don’t want to undress in front of men but I have no choice.*  **Patients’ perceptions of the seclusion**  *Sometimes you’re hungry, they don’t open the door, you want to go to the bathroom, they [the staff] don’t open the door. You think that’s normal. When we urinate on the floor, they come and open the door and tell you to go get the mop and clean up the mess. Why when I hurt myself, they don’t come and ask me what’s wrong?*  **Coping strategies while in the seclusion room**  *In the isolation room, it happens that I cry, I bang on the door, I’m like that...I take my eyeglasses, I break them. I said to myself, if I am often like this, they will see, they will see that there is something wrong with me...I want to talk.* |
| Holmes et al 2015  Canada | To explore the lived experience of seclusion in a forensic psychiatric hospital | Interpretative phenomenological analysis, semi structured interviews, content analysis | During admission, within six months of seclusion | Forensic psychiatric hospital | 13 forensic psychiatric inpatients who had experienced seclusion within the last six months | **Experiencing seclusion**  *It’s very negative. You feel really bad when you’re about to go in there and when you’re in there, because you have nothing to do. You just lay there for hours, not being able to talk to anybody. It’s very overwhelming.*  **Assessing quality of care**  *Whenever I wanted to speak to the staff, I’d have to bang on the door or yell. And then, I never really knew if they were going to come or not.*  *I hate it...they take away your clothes, and… everything you usually have like a nightcap… shampoo and all that. So it’s more like a police station cell.*  **Space of confinement**  *They’re [anti-rip clothing] like [a] potato sack. Once you’re naked, you put it on. It only goes like past your bottom and it’s very uncomfortable. It’s like a crazy suit almost.* |
| Hughes et al 2009  UK | To explore patient perceptions of the impact of involuntary inpatient care on self, relationships and recovery | Semi structured interviews, thematic analysis | 11 participants were interviewed following discharge. One was inpatient under section. | Unclear | 12 participants with previous involuntary inpatient admissions. 11 were outpatient, one was inpatient and detained under section  Five male and seven female, age range 19-62, range of diagnoses and number of admissions | **Views of self**  *I had no self-respect when I left there whatsoever.*  **Experience of relationships and interactions**  *They used to take away my furniture, so I was left with a mattress on the floor. And, no sheets, no bedding [. . .] So those were the ways that they used to punish me.*  **Medication**  *I couldn’t talk, my mouth was locking, my mouth was like twisting, it really hurt [. . .] And I couldn’t talk to let them know what was wrong.*  *They took me back to the room, they put me face down on the bed, actually holding my face into the cushions, so that I couldn’t breathe. I was fighting and fighting. And they were saying, um, go on, pull her trousers down and stick it in her arse. I thought they were raping me.* |
| Insua‐Summerhays et al 2018  UK | To explore the perspectives of staff and patients on factors which facilitate or inhibit therapeutic engagement during 1-1 observations | Critical realist epistemological framework, semi structured interviews, thematic analysis | During admission | Two acute adult psychiatric wards | 28 patients participated. Participants had been on 1-1 observations within the past year.  Wide range of ethnicities, ages, diagnoses, reason for observation and length of time on observations | **An uncomfortable silence**  *I feel like they don’t really care and they’re just here to do a job… That irritates me. They don’t really pay attention to you or they don’t look at you or talk to you, like they don’t really want to know.*  **Feeling judged and misunderstood**  *They look down at you and they start kissing their teeth and stuff… they’re meant to be the staff, so they should be being more mature enough to have a conversation and ask you why you’re upset … Instead of judging you and giving you dirty looks.* |
| Isobel 2018  Australia | To explore voluntary and involuntary service users’ experiences of care during hospitalisation | Mixed methods questionnaire, structured interviews, mixed inductive-deductive descriptive analytical approach, content analysis, thematic analysis | During admission | Two acute inpatient units | 67 participants (40 involuntary and 27 voluntary)  No other demographics available | **Interactions with staff**  No quotes presented.  **Involvement in care**  No quotes presented.  **Ward environment**  No quotes presented.  **Treatment**  No quotes presented.  **Scared to express yourself**  *I feel safer here. But I feel a bit like a specimen that’s being viewed. I can’t express stress or concern. Even about things that should be stressful.*  **I feel powerless**  *I’m not here by choice. I can say what I think but I don’t have the final say and there’s not much I can do.The way I behave is the only thing I can control.*  **Sometimes I feel like I am on the outside looking in**  No quotes presented. |
| Jeffs et al 2011  Canada | To explore how service providers and users experience and define near misses | Exploratory qualitative design, semi structured interviews and focus groups, content analysis | During admission | Three inpatient mental health units | 28 patient participants  No demographics available | **Fearing harm and feeling threatened**  *In an environment like this, I always constantly feel threatened. You feel helpless all the time. If I just walk down the hallways is a near miss situation for me.*  **Delay or error in care due to lack of communication across transition points**  *He [psychiatrist] forgot to put on the order drug x that really takes a lot of anxiety away from me. But three days went by and I could snap.* |
| Johansson and Lundman 2002  Sweden | To illuminate the experience of being subjected to involuntary psychiatric care | Phenomenological hermeneutic method  Narrative interviews | Within two years of discharge from an involuntary admission | Patients own home and mental health centre, at patient choice | Five patients, (three female and two male)  Age range 27-49  Involuntarily detained within the last two years | **Being restricted in autonomy**  *I fought, I kicked against, I put up my feet because they carried me by my arms, so I put up my feet against the door so they could not drag me further away.*  **Being violated by intrusion on physical integrity and human value**  *I become aggressive when they use violence, it’s an encroachment when they don’t say anything but just catch hold of you and drag to the bed and give the injection with force and a lot of people are holding you. They didn’t have to use violence. They didn’t need a whole army from two wards.*  **Being outside and not seen or heard**  *[About the civil court] . . . I don’t understand why you have to participate in that, I don’t understand a thing about it anyway. I mean, I can’t defend myself there. Butit is a formality you have to, I suppose.* |
| Johnson et al 2004  UK | To investigate women’s experiences of admission to women-only crisis houses and to general acute wards | Semi structured interviews, content analysis | Within three months of discharge | Participants own homes | 30 women admitted to women-only crisis houses  20 women admitted to three different acute psychiatric unit, voluntarily | **Safety and fear of other service users**  *And I’ve had bad experiences, yes . . . a male patient getting into bed with me at night and that sort of thing.*  **Effects of the crisis house and hospital environment**  *Going into hospital is such an undignified and degrading, horrible feeling. And in many cases it makes it worse and took me a lot longer to recover.*  **The stigma of admission**  *To being hospitalised for mental illness, which is kind of like a stage further down, going to the mad house.*  **Contact with staff and opportunities to talk**  *I didn’t get much attention really. They didn’t seem to ask many questions or try and help me particularly. It seemed almost like they were just supervising basically.*  **Involvement in care**  No quotes presented.  **Management of medication**  *The decision was completely out of my hands, that’s why I refused taking medication at the beginning . . . I hadn’t seen a proper doctor who could tell me what’s wrong with me. I’d been told [what to take] and I didn’t like that, I didn’t respect that, I felt there was no respect for me from the professionals who prescribed me the medication.*  **Other treatment and activities**  *There were no activities, just carpentry and a bit of clay work and if you’re not artistic or anything, you just can’t do it. There was nothing to do but mope around in bed all the time and sort of sleep.*  **Women with children**  *[We needed] somewhere we could have gone, so that he [her child] was out of the main acute hospital environment. Somewhere for us to go that was quieter. There was nothing offered from the hospital.*  **Assessment and admission**  *It [pre-admission assessment] is an upsetting procedure when you are very vulnerable, very upset and very, very low in yourself.* |
| Kalagi et al 2018  Germany | To explore the opinions and values of staff and service users in relation to open door policies in psychiatric hospitals | Semi structured interviews, content analysis | Some patients were interviewed while still inpatient. Others were outpatient at time of interview | Hospital setting | 15 patients, both inpatient and outpatient  12 male and three female | **Seclusion**  *Hence the rest room, the padded room. So they can go in there and let off steam without end and when they’re calm again, they can come out again; instead of being mechanically restrained. Because when you getmechanically restrained, it rather causes more frustration.*  **Increased freedom of movement and outdoor activities**  *The problem was that I had to walk all the time because, due to the antipsychotics I got, I had restless legs symptoms. So somehow walk all the time, and so I constantly walked in a circle in this courtyard garden and was annoyed that I couldn’t get further out.* |
| Katsakou et al 2011  UK | To investigate perceptions of coercion of admission among legally voluntary patients | Exploratory mixed methods study, questionnaire, semi structured interviews | Within three months of admission | Both in hospital and participants homes | 36 patients, 23 of whom reported coercion | **Hospital treatment not effective/need for alternative treatment**  *The hospital is not good for me, it makes me more stressed. I like to be in me own flat and go to the day hospital everyday...that's where you get more respect, cause here [in hospital] they think you are an animal.*  **Not participating sufficiently in the admission and treatment process**  *I didn't really decide, they decided for me...I thought that if I didn't say yes then I would be sectioned, so really I did feel coerced...it certainly didn't feel like I had a choice, so I got angry.*  **Not feeling respected/cared for**  *There's a whole team there and they don't listen to you; they TELL you...it just made me feel like I wasn't human and nobody actually took my point of view into consideration.* |
| Katsakou et al 2012  UK | To explore involuntary patients’ retrospective views on their perception of hospitalisation | Grounded theory, semi structured interviews, thematic analysis | Between three months to one year after index admission. All had been discharged at point of interview | In patients own homes | 59 involuntary patients admitted to acute wards | **Need for non-coercive treatment**  *There weren’t any other patients that self-harmed and I don’t know if the hospital understands. I need somewhere that’s going to help me, and understand, and work with me through it, not force medication on me.*  **Unjust infringement of autonomy**  *Once you’ve been in hospital if they say you’ve got to go into hospital, you have got to go; like being under the surgeon’s knife: once under the surgeon’s knife, always under the surgeon’s knife.* |
| Khatib et al 2018  Israel | To explore peoples retroactive accounts of their experience of restriction and restraint | Semi structured interviews | Following discharge | In the community | 15 participants, all under community care following involuntary admission having been restrained | **Duration of restriction**  *After being tied I received an injection and fell into a deep sleep. When I woke up two hours later...I thought that I would be untied, but I was still tied to bed. I remember myself calling the nurse and begging her to untie me...She answered ‘no! Not yet!’.*  **Lapse of time from the time of restriction to first contact with a staff member**  *The nightmare started once the door was closed and I was tied to bed and alone. I was terrified...I thought I was about to die, I screamed and begged not to be left alone.*  **Accusing interactions**  *‘You are continuing to run wild even though we tied you.... This is not right...you must control your behavior...you and only you are responsible for this...why did you act so aggressively to that nurse? You don't under-stand a thing if you choose to act like this’...And then she left the room without even waiting for me to respond...I felt I couldn't trust her...She had no idea what impact her words had on me…* |
| Kim et al 2007  USA | To explore the use of psychiatric advance directives for patients with serious mental illness | Semi structured interviews | Following discharge | Community based treatment program | 28 participants under community based treatment programs, following discharge from inpatient psychiatric services | **Difficulties communicating PADs [psychiatric advance directives] to inpatient staff**  *They wasn’t really listening to nothing that I had to say, they weren’t really paying me no attention, it was like I wasn’t really there, I was just there to be admitted and I didn’t have no say–so about nothing that I showed anyone. I didn’t want to end up with them putting me in seclusion or something so I just went along with whatever they said. So I didn’t want to get in no trouble.* |
| Klingemann et al 2022  Italy, Poland,  UK | To explore different forms of treatment pressures put on patients during admission to psychiatric hospitals | Semi structured interviews, thematic analysis | Unclear | Unclear | 108 patients with inpatient experience  Range of diagnoses and demographics | **Treatment pressures: Persuasion**  *The doctor suggested that it would be better to have me admitted, and I accepted it, though I really didn’t want to. I tried [to say that to the doctor] but I wasn’t feeling well, and the doctor explained that it would be better for me, so she persuaded me, and even called my mother.*  **Treatment pressures: Interpersonal leverage**  *Well I knew I was unwell but I didn’t want to go to hospital, so my brother, mainly my brother was in charge, yeah my brother took me in the hospital and my family, my sisters all ganged up.*  **Informal coercion: Threat**  *She warned me that if I didn’t go to the hospital, they’d take me by force…I had no choice.*  *It was very difficult for me to accept that it was so easy for my family to put me in the closed ward.*  **Informal coercion: Violence**  *I remember (regarding being admitted). They had to push me into the car by force [bitterly] – that was mother, my aunt and my goddaughter. I didn’t want to, so they were going to call an ambulance. I ran away at the hospital claiming I was healthy. And they chased me and held me down [bitterly].* |
| Klingemann et al 2020  Belgium, Germany, Italy, Poland,  UK | To explore the experience of patients and clinicians regarding specialisation and personal continuity of care | Semi structured interviews, thematic analysis | Unclear | Unclear | 188 patients, maximum variation sampling, with experience of receiving specialised care or receiving personal continuity of care | **Negative experiences with personal continuity of care approach**  *They didn’t have the time… we’re talking about the bare minimum.*  **Negative experiences with a specialised care approach**  *It is often very strenuous, if one has to keep on starting from the beginning and has the feeling ‘I’ve just told someone else everything’.* |
| Kogstad 2009  Norway | To investigate violations of dignity from a patient perspective | Narrative reviews, qualitative content analysis | In the community following discharge | Remotely written and posted to researcher | 267 narrative reviews | **No treatment apart from medicinal treatment**  *I was forcibly sent to the hospital because I said I felt like committing suicide. I was heavily medicated and had only one talk with the doctor during my entire stay. I felt I was left totally on my own together with other patients who screamed and smashed furniture. I shared a room with people who scared me. It was a painful experience.*  **Trauma experiences disregarded**  *Once when I was in my thirties, I was in the hospital. The anxiety came back and I asked if I could talk to a psychiatrist. I thought that at last I would be able to open up and talk about the incest I had experienced as a child. His answer was: It was such a long time ago and should just be forgotten.*  **Children/family not cared about when person is committed**  *I was committed and had to leave my children, aged 2–19 years. No help was offered. I was neither listened to nor taken seriously, and did not get any help from the community health services. I was just given medicines with painful side effects.*  **Commitment**  *This strange doctor concluded that I should be sent to the hospital. I objected and said: “It will not help.” But a person in my situation suddenly has no right to express herself. The doctor said: “Then it is a compulsory admission!” I objected and objected, but my voice did not count any longer. My husband signed the paper (after the doctor threatened that if he didn't, the police would do so). I don't think I have ever felt so deceived before. I was angry, sad and empty (…), and overwhelmed by the feeling of being totally turned down. I had lost everything. It felt like mental rape.*  **Forced medication**  *I was medicated by depot injection, but the way they did it was wrong. I didn't want the medicine. Four-five people were in the room. One gave the injection, while the others held me. I resisted. I was afraid. After this, they all left. I was alone. Later, I didn't want contact with the staff at all. I hid under the bedspread.*  **Punishment**  *I was confined and did not want to get out of bed. I was punished with no more cigarettes. They took away all I had and locked me into a room for three days.* |
| Kontio et al 2011  Finland | To explore inpatient psychiatric experiences of seclusion and restraint | Focused interviews, inductive content analysis | Between two and seven days after seclusion or restraint occurred | Six acute inpatient wards | 30 patients (11 female and 19 male)  Age range 20-64  Range of diagnoses | **Patients’ experiences before seclusion/restraint**  *I didn’t understand why they put me into the seclusion room and I never got information on this. The staff was reluctant to provide information on why and how long, what next.*  **Patients’ experiences during seclusion/restraint**  *I felt fear and anger, especially toward those who put me into the seclusion room. Nurses and physicians used power and authority over patients. I didn’t know where I was and how long it lasted, it was terrible.*  **Patients’ experiences after seclusion/restraint**  *There was no chance to talk about my experience.* |
| Kuosmanen et al 2007  Finland | To explore whether patients experience deprivation of liberty during psychiatric hospitalisation and to explore their views on this | Explorative design, semi structured interviews, inductive content analysis | Following discharge | In the community | 51 patients recently discharged from two acute psychiatric wards | **Restrictions on leaving the ward**  *…at the beginning everything was restricted, it was like being in prison.*  **Restrictions on communication**  *They took away my cellphone and I was allowed to use it only at certain times.*  Coercive measures  *I was so fuddled by drugs… three men came and they stuck a needle in my ass.*  **Confiscation of property**  *They took my clothes.*  **Patients’ feelings about their deprivation of liberty**  *It would be good to hear the justifications; it is hard when you have to interpret everything alone; I need to talk this through.* |
| Lamanna et al 2016  Canada | To explore inpatient and clinician perspectives on factors affecting verbal and physical aggression by psychiatric inpatients | Interpretive theoretical framework, semi structured interviews, inductive thematic analysis | Between one day prior to discharge, up to 14 days after discharge | In the community | 14 inpatients (nine female and five female)  Age range 18-77  Range of diagnoses, length of admission and number of previous admissions | **Personal factors: Major life stressors**  *The whole day was just devoted to being upset because of my kids: I didn’t know where they were.*  **Personal factors: Experience of illness**  *I was upset, because it’s telling me to follow, but I don’t want to follow...the voice [said] to do this, do that, but Ido not want to follow, so I just throw things.*  **Personal factors: Interpersonal connections with clinicians**  *That’s what pisses me off, is that some people...don’t really have the time of day for you, and they just close theglass like they’re mightier than thou.*  **Organizational factors: Physical confinement**  *You’re trying to put a gorilla in a small, you know – like a lion in a small [forms box with hands]– it’s crazy. It makes you go crazy...I start kicking the door and trying to break it open.*  **Organizational factors: Behavioral restrictions**  *It’s about freaking out over an item that I can’t have, because they took away my freedom.*  **Organizational factors: Lack of engagement with clinicians and treatment decisions**  *She didn’t tell me what kind of pills or nothing, she just said ‘take the pills, you have to take these pills’. I said ‘I’m not taking any pills’, so she called the security guard...I’m not going to take something I don’t know.* |
| Larsen and Terkelsen 2014  Norway | To explore how patients and staff experience coercion in a locked psychiatric ward | Ethnographic fieldwork, interviews (staff only), phenomenological approach | During admission | Inpatient acute psychiatric ward | 12 patients (nine male and three female) | **Corrections and house rules**  *The first offense was on arrival when Jack (E) stood in front of me in the hallway, arms crossed, explaining the rules. He looked as if he was working as a doorman outside a disco. He explained the rules here in a top-down manner.*  **Coercion is perceived as necessary**  *It implied accepting involuntary admission voluntarily, so they could force me to take antipsychotic medication.’*  **Significance of material surroundings**  *Hell is the right word. When you don’t understand why, it’s like going through a real Hell.*  **Being treated as a human being**  *Most of all they are concerned about my head. They want to give me medication for hypomania, but [...] I’m not concerned about the diagnosis. My opinion is that we are complex [...] body and soul, but here the only thing that counts is the head.* |
| Lawrence et al 2019  USA | To explore the impact of coercion on treatment alliance from the patient perspective | Semi structured interviews, grounded theory | During admission | Two psychiatric wards | 50 patients (31 male and 19 female)  Range of ethnicities and diagnoses | **Submitting a sign-out letter**  *The conversation [about discharge] needed to happen and it wasn’t happening, and I needed to feel more in control of what was going on.*  **Involuntary hospitalizations**  *I lost trust in psychiatry completely.*  **Locked doors**  *Yeah. I feel like, they have so much power over me, and I really just don’t like it. I feel like it’s more of a power thing than it is, like, a compassionate ‘they’re trying to help me’ kind of thing. I just feel, I definitely feel trapped and I feel powerless. And like they are the ones; they are the ones that are not allowing me to leave those locked doors. So, yeah, I do think that that affects it.* |
| Lilja and Hellzen 2008  Sweden | To explore former psychiatric inpatients’ experience of their admission to psychiatric inpatient units | Qualitative, semi structured interviews, content analysis | Following discharge | In the community | 10 patients (three males and seven females)  Inpatient within five years prior to interview | **Being seen as a disease**  *Even if the premises are new the content is the same as it always has been, that is: loneliness.*  **Striving for a sense of control in an alienating and frightening context**  *One has to struggle for one’s identity.*  *…* *there should be different levels in hell/.../here they mix healthy patients with severely ill ones...living together under they* [sic] *same roof isn’t easy.*  **Meeting an omniscient master**  *.. . . usually you get pills, not someone who listens to what you have to say/.../I think you are fed with pills...it’s important that you have a physician who talks...a physician who listens to you.* |
| Lindgren et al 2015  Sweden | To describe the features of psychiatric inpatient care for women who self-harm | Participant observations and informal interviews, content analysis | During admission | Two acute psychiatric wards | Six female patients with history of self-harm  Age range 21-37  Three voluntary and three involuntary | **Confusing environment**  [Observation] *Paula and Amanda* [patients] *go to the office door, knock and say through the locked door, ‘There is blood on the floor again, could you please clean it up? We find it disgusting’. The nurse comes out into the corridor and begins to clean the floor while her colleagues go to the patient and say, ‘We think it is time you rest your feet now, let us follow you to your room’. Amanda, Paula and the other patients witnessed the nursing staff having to force the patient into his room.*  **Routines and rules lacking consistency**  *The patients were prepared to go for a smoking break and waited next to the atrium door according to routines and the information they had got. The nursing staff were not there to open the door and t*h*e patients were eager to smoke. Lisa, ‘Where is the nursing staff? I want to smoke’. The atmosphere was tense and the patients began to complain to each other that although the nursing staff insisted that patients must be on time, they (staff) did not arrive on time.*  **Waiting in loneliness**  *No one else was available to talk to her and no one took notice of her frustration. Amanda, who overheard Paula’s discussions with the nursing staff, responded to her and said, ‘It is useless having nursing staff here when they don’t talk to you’.* |
| Lindgren et al 2004  Sweden | To describe how people who self-harm experience received care and their desired care | Personal narratives, content analysis | In the community. Five of the participants had been discharged within the last year | Community mental health service and psychiatric clinic | Nine females with inpatient and outpatient psychiatric care experience | **Not being seen as human does not confirm self**  *But there I got treated totally . . . like a car that had been sent for repair. It was as if I didn’t even have a soul. It was shocking, really awful.*  **Being stigmatized does not confirm humanness**  *It seems like staff often mean ‘You are a borderline . . . , so it is either this or that way, there is no use in giving you treatment because you will resist it’, then I feel labelled.*  **Being disconnected from staff/environment does not confirm self**  *I have felt that before: you are never as lonely as when you are an inpatient in psychiatric care.*  **Being doubted by staff, and having unmet expectations does not confirm self**  *W*h*en those that are supposed to help doubt me, then I do not believe in myself.*  **Not being understood by staff does not confirm self**  *I feel so sorry for the staff, it shows that they don’t understand that it is a trauma to be locked in one’s self.* |
| Lindkvist et al 2021  Sweden | To gain knowledge of the meaning of brief admission for self-harming individuals at high risk of suicide with histories of extensive psychiatric inpatient care | Phenomenological hermeneutic method, semi structured interviews | Following discharge | Participants own home or mental health clinic, as chosen by participant | Seven patients with more than 180 days of inpatient care in the previous year | **Being worthy**  *I went to the psychiatric emergency unit and said, “I am going to take pills today and I need help not to”. And then he said “well, you may begin in day-care in two weeks.” And I had brought a bottle of/promethazine/because I knew that’s what they were going to say. I took the pills. And then he changed his mind. The healthcare system has always been that way, necessitating self-harm. I mean, if you can’t explain—and I can’t explain—you have to self-harm to get help.*  **Struggling in early help-seeking**  *Like, “listen to me”. They are ignoring me saying that I can handle it. It is, in a way, both condescending and disabling, or perhaps not disabling, but you do feel a bit ridiculed. And not taken seriously.* |
| Ling et al 2015  USA | To examine debriefing data to understand patient experiences of restraint | Audit of inpatient debriefs (qualitative data analysed), thematic analysis | During admission, debrief post incident | Acute inpatient wards | 55 patients | **Antecedents to restraint events: Lost autonomy**  *I freaked out about having to stay in the hospital for a week or two with nothing to do.*  **Antecedents to restraint events: Interpersonal tension**  *I tried to do damage to a person, because he was making fun of me.*  **Antecedents to restraint events: Feeling unheard**  *I was telling staff that I did not need any restraint and expressed full control all along. If the staff were empathetic enough they may have understood that there was no need for forced restraints/medications.*  **During the restraint events: Fear and rejection**  *Hurt, frightened, made me feel like prey. Feel like somebody is going to cut me into pieces. I don’t want to come back.*  **Post restraint events: Lost trust**  *I felt there is no rationale behind many of the decisions made towards me.* |
| Longo and Scior 2004  UK | To explore how individuals with intellectual disabilities and their carers experience psychiatric inpatient admission | Semi structured interviews, interpretative phenomenological analysis | Admission within the last 12 months, regardless of whether still inpatient or not | Seven psychiatric wards across three hospitals | 29 patient participants | **Lack of control**  *I didn’t quite understand it* [CPA meeting]*. I’ll be going somewhere else. I am scared. I don’t want to go there. I’d like some more time here. It’s too late now. They have decided. The doctors and the nurses.*  **Protection and nurture versus indifference and harm**  *You try to speak to them but they ignore you. They keep saying they’re busy.*  **Negative aspects of the environment**  *Dirty.*  *Too hot.*  *Closed in.* |
| Lorem et al 2014  Norway | To discuss the patient’s moral evaluation of coercion in mental health care | Qualitative study, participant observation, semi structured interviews, thematic analysis | Four interviews took place during admission, the final interview following discharge | Acute psychiatric ward  One interview in patients own home following discharge | Patient observation over 213 hours (number of patients observed unknown)  Five patient interviews | **Fighting or resisting**  *They confiscated everything. On the one ward, I was allowed to have some personal things. They began to confiscate more and more. I had hardly any objects in the room.*  **Resignation**  *So you get very distressed when you’re admitted against your will . . . because you’re keen to be discharged, and sodo all it takes for you to get out. Anyway, I feel like a tiger in a cage.* |
| Lu et al 2017  USA | To evaluate the subjective experience of psychotic symptoms and treatment in patients with multiple episodes of psychosis | Grounded theory, semi structured interviews | During admission | Inpatient psychiatric unit | 63 patients (36 male and 27 female)  Range of ethnicities, diagnoses, traumatic event types, and number of previous admissions | **Feeling confused about a long stay in the hospital**  *Since 2001 only released for 4 months and in the hospital for over 6 years.*  **Coercive treatment**  *The fact that I was restrained, three guys restrained me [and] gave me a med.*  **Forced medication**  *Having them give me Risperdal every two weeks; When I was in the quiet room, they told me if I don’t take it, then they will force me to take it—restrain me to the bed.*  **Mistreatment**  *Verbal abuse by staff [was the most traumatic aspect of treatment]. They make fun of our feelings, make me behave the way I don’t want to. If I don’t keep staff happy they will drop my level like a punishment.*  **Being with other psychiatric patients**  *Seeing a patient pulling her hair and throwing up was the most traumatic.* |
| Maloret and Scott 2017  UK | To explore how mental health inpatients with autistic spectrum conditions experience and cope with anxiety when admitted to an acute mental health inpatient ward | Qualitative naturalistic research design, semi structured interviews, interpretative phenomenological analysis | Following discharge | Participants chose location, interviews occurred in own homes, day centres and colleges | 20 participants with two or more weeks psychiatric inpatient experience.  All had diagnosis of an autistic spectrum condition | **Anxiety**  No quotes presented.  **Fear**  *I remember the fear and anxiety within me and how it pushed me into a rage. I felt very paranoid about the peo-ple around me and considered that it was them against me.*  **Lack of routine and structure**  *After the first four to 5days of my admission period, my stress levels began to decrease as I began to understand what I was there for and how I should spend my time.*  **Sensory profile of the unit**  *I had problems with my personal hygiene on the unit be-cause I did not like the water in the shower, it fell harder than the water I had at home and it was actually quite painful to have a shower or bath or even wash, so much of the time I chose not to.*  **Food**  *Additionally, I had problems with the food. I didn’t like the texture or the taste and certainly not the smell of some of the foods that came from the hospital kitchen.*  **Isolation**  *On the unit finding a quiet area was impossible, even the areas which were called ‘quiet areas’ were always quite busy and I can never find time just to be on my own. There was four beds in my dorm ...even with the curtain round my bed I was never alone. I found this unbelievably difficult to deal with and as a consequence I spend my entire time on the units on the edge of my nerves.*  **Stopped eating**  *I was feeling so worried all of the time, I simply didn’t have an appetite.*  **Self-harming**  [Aim was to] *knock the fear and anxiety out of his head.* |
| Mayers et al 2010  South Africa | To explore the perceptions and experience of patients exposed to sedation, seclusion and restraint | Focus group, semi structured interviews, thematic analysis, questionnaire | Unclear | Unclear | Eight patients participated in initial focus group  43 patients were interviewed | **Inadequate communication: Service-provider-service user communication**  No quotes presented.  **Inadequate communication: Isolation**  No quotes presented.  **A violation of rights: The use of seclusion as punishment**  *In hospital when I still getting frustrated asking for medication ... the night sister she told me that I must come out, I must come out, she’s going to give me some medication, and you know what she did to me? She put me in seclusion.*  **A violation of rights: Excessive/inappropriate use of force**  *Treated as animals and not as human beings in distress. They took my hands like this, you know, and the one put their knee on my back in my kidneys and they pulled me … we’re also human beings, we are not animals.*  **A violation of rights: Lack of respect for basic human dignity**  *I would come up with something that will please them otherwise if I don’t please them, I know he’s going to increase that haloperidol ...he would ask me ...what can you sing for us? Even if you haven’t got a song, you have to compose your so.*  **Experience of distress**  *It wasn’t nice for me... I wouldn’t like to be in such a situation because they don’t treat the people very nicely ... it’s not a nice effect ... and I wouldn’t like to be hospitalized again.* |
| McBride et al 2014  UK | To explore the experience of high-level observations | Mixed methods  Questionnaire, focus groups and 1-1 interviews | During hospital admission | High secure hospital, four participating wards | 30 semi structured interviews with patients.  Focus group one had 20 participants. Focus group two had 24 participants | “First you hate it, then you get used to it, then you like it and then you depend on it.”  “I was on them for too long”  “I hated being watched all the time, it was like big brother” “I can’t sleep because my door’s open and people are watching me at night”  “They were pointless, no-one told me why I was on them and I could have self-harmed if I wanted to anyway”  Most patients felt their privacy was being breached |
| McGuinness and Dowling (2012)  Ireland | To explore the impact of and experience of involuntary admission | Interpretative phenomenological analysis  Semi-structured interviews | Mental health centre | In the days prior to planned discharge | Six participants who had been detained, though about to be discharged | **The early days**  *The one main thing for me is that nobody ever had a clue how long it was going to take, how long it was going to be before I got out, you know, there was no exit.*  **Experience of treatment**  *It wasn’t needed like, the injection, I would of actually taken them. I remember saying . . . them saying “do you refuse to take it orally?” but I didn’t hear them properly and I just didn’t know what to say and I was panicking, I said yeah I do refuse, I said yeah, and then I realised straight away and then the needle was in.*  **Moving on?**  *I was just f*** angry but I wasn’t allowed show it, you know there’s no sort of place to show anger . . . that means there is something wrong with you, but it doesn’t mean there anything wrong with you it means your normal but if you get angry it’s like “he’s angry” lets calm him down.* |
| McGuinness et al 2018  Ireland | To explore individuals’ experiences over the course of an involuntary admission | Semi structured interviews, grounded theory | Three months following discharge | In the community | 50 patients who had been involuntarily detained | **Losing control: Feeling violated**  *They [assisted admission team] just dragged me … They put me against the floor, used violence … they handcuffed me and they put me in this plastic yellow blanket and put me in a van or something … I didn’t know where I was going.*  **Losing control: Being confined**  *They [staff] wouldn’t let you out in case you ran off … and not being allowed to get out and have fresh air was a major factor to me … I felt restricted … If you think you’re in a prison, you’re not going to get much better.*  **Regaining control: Resisting the system**  *I was trying to break free...I was so shocked and angry...I was like shouting and all that.*  **Maintaining control**  *It made me aware of how vulnerable I am the system that’s there … it’s very controlling.* |
| Meehan et al 2000  Australia | To explore how patients receiving acute inpatient treatment describe and construct meaning about their seclusion experience | Naturalistic qualitative design, semi structured interviews, thematic analysis | Secluded within the seven days prior to interview | Two acute inpatient wards | 12 patients who had experienced seclusion in the previous seven days | **Use of seclusion**  *I was hauled back here and placed in seclusion. Policemen dragged me out of the house, even though I was offering no resistance, and then I was stripped and placed in seclusion. Yes. Quite barbaric is what I thought of it.*  **Emotional impact**  *By the time I was out I didn't dare talk to anyone or do anything, you know, cause I was frightened I'd go back in.*  **Staff-patient interaction**  *I don't understand why they felt the need to put me there. I still don't understand that, and no one will discuss it with me. They could have sat me down and explained why I'd been through all that hell.* |
| Molin et al 2016  Sweden | To explore the patient experience of everyday life in psychiatric inpatient care | Grounded theory, semi structured interviews | Six were inpatient at time of interview, 10 were in the community following discharge | Mental health clinic | 16 participants, (14 female and two male)  Age range 20-51  Range of diagnoses with multiple previous admissions | **Staff makes the difference: Adapting to absence of interaction with staff**  *I think it is a big problem that you never see the staff. They sit inside the office all day. You have to stand and knock for a long time if you want to reach them.*  **Looking for shelter in a stigmatising environment: Adapting to a destructive environment**  *When you get to the ward, there is zero stimuli. There is not a single curtain, and there are only three chairs that are screwed into the wall in the hallway. There are no bedside tables. There is nothing. You only get a feeling that you should not be here.*  **Facing a confusing care content**  *One day is very irregular and it differs from day to day. It is very, what should you say ... a bit foggy. There is no real knowledge of the patients.*  **Adapting to an unclear structure**  *One day is very irregular and it differs from day to day. It is very, what should you say...a bit foggy. There is no real knowledge of the patients.* |
| Mottershead et al 2020  UK | To explore patient experience of forensic and non-forensic services | Mixed methods, survey, chi squared tests, content analysis | Unclear | Surveys completed remotely | 906 patients who completed Service User Care Experience survey | **Staff**  I don’t feel included staff/patient divide. Because when moved from Emerald no discussion as to why move was happening.  **Quality of life**  *Dignity–once when I was unwell I stripped down naked and then held me in seclusion when I was naked I think it was wrong.* |
| Murphy et al 2017a  Ireland | To explore the experiences of patients admitted involuntarily to psychiatric hospitals | Qualitative descriptive study, semi structured interviews, inductive thematic analysis | Three months following revocation of the involuntary admission order | In the community | 50 participants (29 male and 21 female), all had been detained involuntarily  Range of diagnoses | **Feeling trapped and coerced**  *I suppose when you’re involuntary, having that sort of label on you and knowing that you’re trapped there [in the hospital]… feel very much like you’ve had your human rights taken away, you feel imprisoned, and you kind of feel, as I said before, a second-class citizen.*  **Lack of informational and emotional support**  *No [response to question about being provided with information], not really. I did. I knew I was in hospital. I had done the interview [with the psychiatrist]. I wasn’t quite sure why I was there. I was saying, “Why am I here? “You know, because I believed I was fine at the time. I wasn’t quite sure why I was there.*  **Admission induced trauma**  *Leaving the hospital. . . that’s even worse, because that’s when the trauma comes in and the fear comes into your normal life. You have to go to work and keep living this like big trauma caused by these people [involved in involuntary admission experience], and this trauma is the one that’s going to cause more severe and more problems.* |
| Murphy et al 2017b  Ireland | To explore the mental health tribunal experience of people admitted involuntarily under the MHA | Qualitative descriptive study, semi structured interviews, inductive thematic analysis | Three months following revocation of the involuntary admission order | In the community | 23 participants who had experienced a tribunal whilst detained under MHA | **Information provision**  *No, I didn’t get much of it [information]. I just got a booklet [‘Your Guide to the Mental Health Act 2001’,MHC].So, I should have got more information on it, but I didn’t know who to go to, was it the nurses or who to go to.*  **Emotional support**  *They told me, you know, you’re going to have to stay in. We’re making you involuntary. You cannot go home for weekends. You cannot get out, except for being accompanied by a family member. This is ridiculous. I thought this is just something out of the dark ages [result of tribunal]. […] I was just listening to them and I said nothing at that point because I felt this is it now. There’s no point in me saying anything. […] It [the tribunal] was very upsetting because I cried and cried and cried.*  **Inclusive practices**  *I felt [I was] against everyone else because I’m thinking a different way. […] it’s kind of insulting for everyone who is a vulnerable patient just to have those words written down […]…. ‘delusions’,’ lacks insight’ […] and there’s a litany of terms that are listed out, way too flippantly, and everyone nods to each other, because they’re all the same, they’ve all learnt things from the same pages of the book.* |
| Niimura et al 2016  Japan | To explore the challenges faced by patients immediately after discharge from psychiatric inpatient care | Qualitative descriptive study, semi structured interviews, content analysis | Between one- and six-months post discharge | Two psychiatric hospitals, outpatient department | 18 patients post discharge  Eight male and 10 female  All diagnosed with schizophrenia | **Feelings of dissatisfaction being admitted to psychiatric hospital**  *I would not have been so opposed [to the idea of hospitalisation] if it had been a normal internal medicine department. However, a psychiatric ward? That I was sort of against.*  **Inability to find meaning in hospitalization**  *Why do I have to spend all these months in here? [I was opposed to hospitalisation in a psychiatric ward] because I felt that very strongly.*  **Dissatisfaction with treatment in protection rooms**  *It seems that I was acting slightly violently shortly before being hospitalised; however, I have no memory of that because I was not sleeping. My mother seems very upset when she talks about it and so I guess it came as a big shock to her that Iwas bound.*  **Dissatisfaction with inability to speak with staff properly**  *I do not quite understand why it was so good for me to be hospitalized. I feel like it’s as though I’m not in hospital and that it would be the same as if I was staying at a hotel or inn, because I have only spoken to the doctor twice or so.* |
| Norvoll and Pedersen 2018  Norway | To increase understanding of patients’ moral views and considerations regarding coercion | Semi structured focus groups and individual interviews, thematic content analysis | At various points of treatment. Some participants were still inpatient, others had been discharged for upwards of 15 years | Focus groups in the community  Interviews on an inpatient rehabilitation ward | 24 participants (10 female and 14 male)  Age range 22-60  Range of diagnoses and number of admissions | **A need for alternative perspectives and solutions**  *Wrongful use of coercion is connected to the system’s wrongful modelling of treatment*  **Dangers to others: violence and aggression**  *When they are in danger to others’ life or health, then I hold no doubt. I don’t find it mean to put hand-cuffs on someone who is trying to kill another person.*  **Danger or harm to self**  *I think, regarding deprivation of liberty, that it is OK to put someone behind thick walls… But I always put my foot down when it comes to medication. [...] It is so hard for me to accept that someone would put chemicals into me to regulate my behaviour.*  **Paternalism**  *I didn’t want to begin using medication at all. I believed that milieu therapy and such things could help me out ofmy depression, or whatever I had. But my therapist said, ‘No, it’s going to be medication’. And I felt that I had nochoice, really. And I was not even involuntarily committed.*  **The content and consequences of coercion**  *But what I don’t respect is being taken by force to a more or less empty, vacuous, vapid treatment, and chemicals.*  **Discrimination**  *But my life views are pathologised. And that is in fact rather offending. [...] I mean, it’s not approved of, and then you’re crazy. That is a quite provoking statement that is pathogenic in itself.*  **Proportionality**  *Some of the episodes with mechanical restraints that I experienced almost felt like an act of kindness or care.[...] But, most of the situations were different and felt more like a kind of violation, especially because punishment was a significant element.*  **The way coercion is carried out**  *I’ve thought a lot about what characterised those coercive situations that became very dramatic and harsh. What was it that distinguished them from other situations that weren’t like that? And then I remembered individuals who showed me respect, who were nice fellow humans, who didn’t speak to me as if I were an ape from a foreign planet or something like that. But [if staff] merely took me seriously, listened to me, then things were solved in totally different ways.* |
| Ntsaba and Havenga 2007  Lesotho | To explore and describe the seclusion experience | Qualitative descriptive design, semi structured phenomenological interviews, open coding | During admission | Psychiatric hospital inpatient ward | 11 inpatients (four males and seven females)  Age range 20-43  Range of diagnoses | **Psychiatric inpatients’ experience of being in a prison**  *When I was secluded it was like I was in a prison. You see, Madame, I was once imprisoned at … because of not having an identification book (passport), we were locked up, eating food and passing stools in the same room, do you hear that?*  **Seclusion experienced as a punishment, which created an environment where human rights violations were experienced**  *You know nurses used to beat me. They slapped and punched me … when I refused to be secluded. They insulted (me) and pushed me in the seclusion room. I cannot mention those insults, they were bad.*  **Personnel factors leading to an experience of not being supported and cared for**  *I would hit hard on the door calling on top of my voice, for nurses to come and help me but they did not come.* |
| Nyttingnes et al 2016  Norway | To explore patient views of mental health services in relation to coercion | Seminars, reflective dialogue approach, thematic analysis | Unclear | Oslo mental health conference | Approximately 100 participants – professionals, patients, family members and carers  Approximately 25 participants were patients/ex-patients | **Expressions of psychiatry as abuse and war**  *It's unbelievably humiliating to be put in belts [mechanical restraints]. Just as bad as Communism and Nazism.*  **Unwanted medical model**  *Psychiatric care frightened the wits out of me, and if I encounter another crisis, it wouldn't even occur to me to seek psychiatric help again. For a long time, I didn't even dare to visit the GP for physical things, out of fear it could lead to another sudden and totally incomprehensible admission.*  **Pressure and coercion to take medication**  *We need to separate the use of physical force, which sometimes is necessary, from forcing chemicals and poison into people, which destroys the brain.*  **Staff disregarding complaints**  *I begged and pleaded for something other than medications, but that was interpreted as lack of insight. That is incredibly humiliating.*  **Minor coercive incidents**  *But there is also a lot of unregulated coercion, directed by household rules. Being searched, having limited access to leave, and a lot of other stuff, is not written in the law.* |
| Olofsson and Jacobsson 2001  Sweden | To describe involuntarily hospitalised patients experiences of coercion and their thought on how to prevent coercion | Descriptive explorative study, narrative interviews, qualitative content analysis | Within three days of discharge from involuntary care | Psychiatric clinic, inpatient setting | 18 involuntary inpatients (12 female and six male)  Age range 19-52 | **Not being respected as a human being: Not being involved in one’s own care**  *Simply because I said I wanted to be discharged they extended my involuntary status. . . if you don’t behave you will not have permission. . . you have to be good and docile.*  **Not being respected as a human being: Receiving care perceived as meaningless and not good**  *I have had medication several times but it never helped me, it made me more confused. I couldn’t recognize myself as I became a robot. All the time they want to fix the symptoms, they really don’t get to the bottom of the problems causing the trouble.*  **Not being respected as a human being: Being an inferior kind of human being**  *You become a nobody, they can do whatever they want with you, although maybe you are a very valuable person being in a crisis.* |
| Olofsson and Norberg 2000  Sweden | To increase the understanding of psychiatric patients’, nurses’ and physicians’ experience of coercion | Descriptive-explorative qualitative study, narratives | Within three days of discharge from involuntary care | Psychiatric clinic, inpatient and outpatient setting | 21 participants, seven of them patients  All involuntarily detained  Five female and two male  Age range 19-38 | No quotes presented. |
| Parkes et al 2015  UK | To explore the male forensic patients’ experience of transition from one medium secure ward to another medium secure ward | Semi structured interviews in two rounds, staff focus group, thematic analysis | During admission  Interviewed six-weeks prior to transfer and six-months post transfer from old to new ward | Forensic hospital, medium secure | Nine forensic patients from two wards  Five participants had mild-moderate learning disability  Age range 24-51 | **Information**  *We don’t have a choice about being here. How would anybody feel about someone coming into your home and telling you what to do?*  **Transition**  *It was just so very difficult for me to communicate just to know what to say or anything . . . I didn’t want to come out with the wrong thing and upsetting them.* |
| Pelto-Piri and Kjellin 2021  Sweden | To explore opportunities and problems in relation to social inclusion for psychiatric inpatients | Qualitative interviews with patients, staff and managers, three focus groups, content analysis | During admission | Three inpatient wards (one adult, addiction and medium secure ward) | 12 patients (four female and eight male)  Age range 23-67 | **Sensible communication between patients and staff**  *That’s it! For goodness sake, you must be allowed to show your feelings. I mean, anger, it’s a feeling; you get angry about something. Yes. I mean, that’s normal […] It doesn’t work in places like this. Then you get put in strap restraints, for heaven’s sake!*  **Negative views of the psychiatric patient**  *Yes, what happens is you feel less worthy because you’re, like, because you’re sick or because it’s an illness. So yes, it didn’t feel like anything was serious. It just became like a big, just a big playground because no one, no one heard.*  **Minimizing coercion, violence, and injuries**  *I have been in some places where they had to wrestle people down, but no...Out in society I have seen a lot of violence, but not in the wards. But sometimes they have to wrestle people down when they’re deep in their psychosis; they have to give them an injection in the buttocks.* |
| Pemberton and Fox 2013  UK | To explore the experience of emotions for inpatients with anorexia nervosa | Semi structured interviews, interpretative phenomenological analysis | During admission | Two inpatient eating disorder units | Eight inpatients with anorexia nervosa | **Difficulties with emotion**  *I thought to myself, ‘I can’t be doing with making enemies with someone ‘the atmosphere was very, very hostile...some and er, my...wisdom thought, ‘I’ll try and paint a little bit of a better picture of me so at least she’s got an appreciation, because she did say, ‘I know nothing about you...’*  **Difficulties in staff-patient relations: Loss of the individual/’Expectations of care’**  *.... a lot of the time you start to feel...especially if you get upset about anything, you’re treated as a walking, talking illness. (pause) You’re not a human being. Everything you say and do or anything you get upset about, it’s the illness, it’s the illness, it’s the illness...*  **Difficulties in staff-patient relations: Need for control**  *...in the end her broken record technique won and I just thought there’s just no point here so I just got on the bed and didn’t speak to her for the rest of the hour....*  **Difficulties in staff-patient relations: Individual differences**  *...your NA’s* [nursing assistants] *which are the people currently doing our hour to hour observations aren’t as well trained as the very few qualified’s that you have on; they don’t have the knowledge necessary to see beyond that...In all frankness, some of them I think are a bit dim..*  **Difficulties in staff-patient relations: ‘Staff’s understanding of emotion’ and ‘Validity of emotion’**  *Virtually every time I get upset...it’s very much a sort of, ‘oh, pull yourself together’ sort of attitude, unless something specific has happened that they feel you have the right to be upset about, like if someone’s died...They accepted I was upset, sort of allowed it to exist*  **Difficulties in staff-patient relations: Rejection/minimization**  *...I was very angry......But it was just like talking toa brick wall. So there is...I don’t see any point at all in taking the emotion further....*  **Difficulties in staff-patient relations: Support and care**  No quotes presented. |
| Pereira et al 2005  Brazil | To explore the lived experience of long-term psychiatric hospitalisation | Nondirective interviews and drawings, theory of social representations | During admission | Long term psychiatric inpatient hospital | Four females, each having continuous admissions for seven, 15, 20 and 50 years | No themes but quotes from participants:  *I stayed in the locked room . . . it was my first suffering, you know. It was difficult to be there; it was very dark. I stayed there all alone for two days and one night—all on my own.*  *I feel there is a lack of patience on the part of those treating us.*  *I received a lot of electroshock, any undesirable behavior resulted in electroshock . . . or it was the isolation chamber. I cried and even yelled when they shut me in the locked room.* |
| Pollock et al 2004  UK | To investigate patient concerns about the provision of medication information on acute inpatient wards | Focus groups, thematic identification and content analysis | Unclear | In the community | 90 participants across 14 focus groups | No quotes presented. |
| Robbins et al 2005  USA | To explore the consumers’ perspective of sanctuary harm in mental health treatment | Qualitative, thematic semi structured interviews | Following discharge, no specific time frame | Day hospital, community based | 27 participants (11 female and 16 male)  Range of diagnoses and number of admissions | **Hospital setting: Threat of physical violence**  *I used to call it the Goon Squad, right. They would be staff members who came to, I guess, quiet down patients . . . they beat the patients to the floor and put their foot on the dude’s neck and held his arm and held him to the floor and then threw him in seclusion. So I was afraid of them.*  **Hospital setting: The rules**  *[The hospital] had paper pajamas. And they are not something you want to sleep in. They itch like crazy. So I tore mine off and put my own pajamas on, so they had me sleep in the hallway. It’s like, just because I switched from something that’s irritating my skin to something that will not, they stick you in the hall . . . It didn’t make no sense.*  **Interactions with staff: Not knowing consumers as individuals**  *Sometimes certain people [other consumers] would be bothering me, and I wouldn’t want to be bothered and whatever. I was trying to tell them, ‘Leave me alone,’ and then I think the staff would think I was trying to cause them trouble, and they would consequent me sometimes versus consequent the person who was actually starting everything…*  **Interactions with staff: Lack of fairness**  *Nothing didn’t happen, but you know, I was locked up. . . I mean, I didn’t talk or doing nothing, and I was locked up.*  **Interactions with staff: Disrespect and humiliation**  *I remember one morning, I woke up and I laid there, but they wanted us to get out of bed real early that morning . . . I didn’t see nothing wrong with it. I didn’t want to bebothered. I wanted to sleep. And one or two of those nurses throwed some cold water on me, and then I got up!* |
| Rojo et al 2009  Spain | To evaluate the level of patient satisfaction at an eating disorder unit | Mixed methodology questionnaire | Questionnaires were posed to participants one month following discharge | Eating disorder inpatient unit | 171 participant (13 male and 158 female)  61 participants had been involuntarily detained | No quotes presented. |
| Rose et al 2015  UK | To explore the perceptions and experiences of service users and nurses in an acute psychiatric ward | Focus groups, inductive analysis | Within two years of previous inpatient admission to an acute psychiatric ward | In the community | 37 service users (16 male and 21 female)  Range of ethnicities and diagnoses  20 had been detained under MHA | **Staff and service users’ interactions on acute wards**  No quotes presented.  **Coercion and control**  No quotes presented. |
| Russo and Rose 2013  Germany, Czech Republic, Finland, Romania, Turkey, Bulgaria, Slovakia, Italy, UK, Greece, Netherlands, Lithuania, Austria, Hungary, Belgium | To explore service user perspectives of human rights in psychiatric institutions | Focus groups, thematic analysis | Unclear | In the community across each participating country | 116 participants in total attended a focus group held in each of the participating countries | **What does it mean to have rights in a psychiatric institution?**  *What kind of human rights can we talk about when people don’t have any rights there? On paper, people might have some rights that are not observed at all (Bulgaria).*  *if you’re admitted to a hospital, you are not able to see your rights [y] I think in general: you have to hand over your right to make decisions to others; you have to take your medication and nothing else (The Netherlands).*  **Psychiatric treatment methods**  *Nothing was done, many people let me down and a real treatment didn’t exist. I have the impression that compulsory admission means that it is all over, no treatment, not really (Austria).*  **Interaction with institutional staff**  *[T]hey worked but they didn’t speak to us (Italy).Close observation has replaced interaction you know, that’s the thing (UK).*  **Quality standards relating to staff skills and psychotropic medication**  *[H]ow to find out how and if the staff has grown out of this professionalism and turned human? [y]Do they hide behind this professionalism and don’t have the courage to bring forth their own humaneness – that should be studied somehow (UK)*  **Limitations of the “Tool Kit” approach to human rights**  *‘Well you’ve got access to medication, you’ve got access a clean bathroom, you’ve got access to all these things.’ But [what] if you are not really treated like a human being, or somebody is not going to sit down and have a real conversation with you [y]? (UK).* |
| Samuellson et al 2000  Sweden | To describe the attempted suicide patient’s perceptions of receiving inpatient psychiatric care | Semi structured interviews, qualitative content analysis | During admission. Between five-80 days after the suicide attempt, though as near to discharge as possible | On hospital ward | 18 patients (13 male and six female) admitted following suicide attempt | **Care and security**  *There was nobody who cared. There were, after all, people in there, and then you didn't feel that you were taken care. I sat there for quite a while, I don't know for how long, but I eventually got up and started walking around a bit. I don't know exactly what I did. Nobody took care of me, at least that's what I thought.*  **Commitment and lack of respect**  *Why do they send me from one place to the other, what is the reason? When I still cannot open up if I don't think that I will get something in return. After each time I met a new person, I thought, `Whom will I meet now', `Will I feel trust for him or her so that I can open up?'.*  **Confirmation and neglect**  *I simply wanted to leave, but then one of the nurses went and locked the door. `You can't keep me here, surely, I must be able to go out if I want to'. No I wasn't allowed to.* |
| Scholes et al 2022  UK | To investigate women’s experiences of restrictive interventions in inpatient mental health services | Semi structured interviews, thematic analysis | 16 were interviewed whilst still detained under MHA | Unclear | 20 participants, from rehabilitation wards, secure services and acute services | **Powerlessness**  *I felt powerless, like I had my legs pinned to the bed, my arms pinned down, my head was pinned down...it just feels like all your choices are taken away from you.*  *I was sexually assaulted when I was a kid, and I don’t like male staff on me ‘cos I feel like they’re gonna assault me again.*  **Dehumanization**  *They are watching me, and they are looking at me like I’m from the zoo. Not a human being.*  *There was a guy and he’s actually been suspended now, he was purposefully hurting me in restraint, bending my wrists. He admitted he gets a buzz out of restraint.*  **Relationships and communication**  *I don’t speak to any of the staff that restrained me, it’s like breaking trust with certain staff.*  *I don’t think they even care to be honest. It’s just a wage packet at the end of the month. Restraint is just something normal to them... they don’t give a fuck about us.* |
| Secker and Harding 2002  UK | To explore psychiatric inpatient experiences for African and African Caribbean service users | Semi structured interviews, content analysis | Unclear | In the community | 26 participants of African or African Caribbean heritage | **Loss of control**  *They could have explained to me exactly what were the problems that they found, um, the symptoms that I was having. And, and . . . but they sectioned me and injected me and shipped me off to a closed ward unit.*  **Experiences of racism**  *I’ve never, in 15 years, I never put my hand on anybody, patient or nurse. Yet they perceive me as being aggressive so I can’t work that quite out. I think it, it’s just to do with black people, you know. It’s like, they don’t, they don’t really. They don’t understand, or like, and do want to suppress black people. That’s my experience. It’s very racist, the nurses, the doctors and that... very racist . . . I think at the time it was appalling what happened to me. If I was white and middle class or something you wouldn’t, they wouldn’t have done that to me.*  **Relationships with staff**  *You know, treated me less than a person. And anyway, so, either racist or just rude because of his job and he probably thought he was better than me – because I was mad.* |
| Sequeira and Halstead 2002  UK | To examine the experience of physical restraint experienced by service users of secure mental health care | Inductive design, semi structured interviews, thematic content analysis | During admission, within 12 hours of restraint | Secure psychiatric hospital | 14 inpatients at a secure hospital  Range of diagnoses  Length of admission ranged from two weeks to over 30 years | **Anger**  *When they restrain you, they hold you down and all that’s going through my head is angry and abused again … it brings back bad memories for me.*  **Anxiety**  *I’m being pinned down that’s what I’m scared of. I panic, it gets worse.*  *It’s scary, and like if they’re restraining you to give you an injection, they’re undoing your trousers or pulling your skirt off. It kind of reminds me of like my past when I was abused and it really gets to you.* |
| Shattell et al 2008  USA | To describe the experience of acute psychiatric care in America | Existential phenomenology, phenomenological interviews | During admission | Inpatient adult psychiatric hospital | 10 patients (four male and six female)  Range of diagnoses | **Imprisoned and confined**  *I like to drink sodas. But ... as of today or yesterday, I don’t think your family members can bring you any more in. But they said we could bring them up from the cafeteria. But they’re like $1.25 [USD]. And that’s just outrageous. And they say, ‘Well, it’s because we had an ant problem.’ Well, how can you bring them up from downstairs? What difference does it make?*  **Like a band-aid on an open wound**  *When you walk out of here ... you walk out on that edge, like that edge you were ready to fall off when they brought you in here. And when you walk out of here, you walk back out on that edge again. So, what’s happened that made it any better? Nothing.* |
| Shields et al 2002  USA | To explore former patients experiences of traditional psychiatric frontline staff in inpatient facilities | Semi structured interviews, thematic analysis | Unclear, post admission however no timeframe specified | Unclear | 18 former patients with inpatient experience | **Not sharing information**  *Before I decide to take all these drugs that are unknown to me, please leave me the med[ication] sheets.” Okay, which is supposed to be your right as a patient, and they said, “No, we’re sorry we can’t do that without a doctor’s consent.” I said, “Yes, you’re required by law to do it.” So they went through the books. They found med sheets for a few things, all of which were out of date, and for some of them, they claimed they couldn’t find the med sheets at all, so I went through the med sheets that they did come up with.*  **Not providing patient agency**  *Well, they didn’t solicit my opinions, wishes, and desires, so respect didn’t come into it. They had no idea what my opinions, [laughter], wishes, and desires were.*  **Being dehumanizing/disrespectful**  *The making you wait to get toilet pa-per when they could just as well get up and get it. It’s just the way they talk to you, the tone of voice, the manner, the attitude. Hard to illustrate but you know it when you see it. Being treated like less than human.*  **Incompetency**  *Someone thought that I was some-one else.... A male nurse walked up to me and said, “Okay, Ms. X,” or whatever, he thought I was someone else, “time for your insulin.” I am not diabetic, he held out an insulin shot and came this close.*  **Escalating situations**  *It felt unnecessary. If they’d just told me “We’ll do this if you don’t stop,” I would have stopped. Sometimes they don’t think before they act. If they’d just said “Stop, or you’ll be taken to jail or restrained or we’ll give you this med,” then I would have immediately stopped.*  **Being apathetic**  *Mostly, I felt at that hospital the staff were indifferent. They weren’t un-friendly or unkind, but they didn’t really go out of their way to build a relationship either. They were marking time, but they weren’t unpleasant.* |
| Sibitz et al 2011  Austria | To establish a typology of coercion perspectives and styles of integration into life stories | Grounded theory, semi structured interviews, thematic content analysis | Post admission, length of time between discharge and interview ranged from six months to nine years | Medical University of Vienna | 15 participants (eight male and seven female)  Age range 32-66  Range of diagnoses | **Perspectives on involuntary admission and coercion: An unnecessary overreaction**  *Five nurses are coming to overpower you,’’ that’s what he said to me, and for thatI blame him . . . I mean, he could have tried to persuade me.*  **Perspectives on involuntary admission and coercion: A practice in need of improvement**  *. . . and then an injection and if you are confused anyway, in my opinion this can make the situation even worse.*  **Integration of experiences into life stories: Over, not to be recalled**  *I don’t want to talk about or remember it, and when I realise in a dialogue that the other person is affected as well and might find it burdensome, then it is even worse...usually I don’t think about it any more because I don’t want to remember, same with regular psychiatric hospitalisations.*  **Integration of experiences into life stories: A life-changing experience**  *It leads to an absolute inferiority complex, I have the feeling that I am not worth talking to other people, already thinking that I am not worth it, well, we can say destroying my personality.*  *At the beginning, after discharge from the psychiatric hospital, I talked a lot about what happened to me but then I changed, I kept quiet about it, because most people can’t deal with it, they actually don’t understand it.*  *Well, I must say, that all that [coercion] impacts the length of treatment, the amount of drugs, then the illness can be pushed, aggravated, the dose of drugs gets higher if you have to deal with injustices in hospital . . . if they don’t believe you then the subsequent depression is deeper, you get into work more slowly, you get into recovery more slowly . . . the traumatic experiences can contribute to a chronic state[of illness].* |
| Strike et al 2006  Canada | To examine how suicidal men use mental health services | Semi structured interview, thematic analysis | Unclear | In the community | 15 males with a history of suicidal behaviour, and substance use disorder/antisocial behaviour/borderline personality disorder | **Disrespectful treatment of patients**  *. . . and the nurses there were mostly belligerent. They didn’t like the people they were dealing with. They thought we were a big load of freaks and they hated their jobs and that was reflected in their treatment of the patients.*  **Insufficient time for proper assessment**  *I don’t find that particularly useful when you’re in hospital and they just make you sit there like a vegetable.*  **Overreliance on medications by physicians**  *When I started feeling anxious or stuff, they’d give me a pill, instead of trying to find out why I’m feeling anxious. If they’re mental health nurses they should be doing more than just giving out pills.*  **Indignities of hospitalization**  *. . when you feel locked up and they take away all your clothes and you’re on a form so you feel like you’re in prison and it’s hard to think I’m getting out tomorrow because you know you’re not and you don’t know when they’re going to let you go . . . I’m in my pyjamas, I take my bracelet off, and I get on the bus to go home for a day, then usually the police show up and bring you back. Yes, it’s like I feel I’m totally locked up and I can’t do anything I want to do.* |
| Sweeney et al 2015  UK | To explore the concept of fear in the lives of adult mental health service users | Grounded theory analysis, focus groups, thematic analysis | Unclear | In the community | 32 participants across four focus groups | **Fear, power and control: Services**  *…they’ve still got some sort of power over you and it’s as if they’re sort of, you know, I feel as though, well I just feel I’ve got to go along with what they say, whether you agree with it or not as a human being, you know, and you should have rights, certain rights.*  **Fear, stigma and discrimination: Services**  *They meet you and they judge you, they stereotype. We all do it, but in that kind of environment it’s detrimental, you know (South European male).*  **Climate of fear: Services**  *You can’t live your life; you can’t be happy one minute and sad the next, angry the next, happy the next – the whole range of emotions that we want to feel as human beings. That thing’s been taken away from us (Black male).* |
| Tan et al 2010  UK | To explore the views of people with anorexia nervosa in respect to compulsory treatment and decision making | Semi structured interviews, thematic analysis | During treatment, eight remained inpatient whilst 21 were discharged and were day patients/outpatients | Inpatient eating disorder unit | 29 patients with anorexia nervosa  Eight remained inpatient at time of interview. The remainder were day patients/outpatients | **Experiences of compulsion in ‘voluntary’ treatment**  *Well, I was given two options really. Either I refused, and they said“if you refuse to put on weight we’re taking you straight up to the hospital and you’re on tubes and drips and everything and, you know, you’re going to have to gain weight or, and you’ll be made an inpatient.*  **Experience of the restrictions of choice in treatment as unhelpful and coercive**  *And also that’s something I find it very hard to talk about, it’s very hard to just say [to the psychiatrist] you know “well actually, you know, you’ve pushed me too far and I’m now throwing up after every meal”. I mean you can’t really say that to someone, it’s very hard to say that, you know. And also I reckon that obviously if I did say that, I just didn’t want to just have to think about the restrictions that would then be placed on me. I’d just be even more restricted.* |
| Thibeault et al 2010  Canada | To understand how patients and staff on acute care psychiatric units experience the unit milieu | Interpretive phenomenology, semi structured interviews | During admission | Acute psychiatric ward | Six patients (four male and two female)  Age range 25-75 | **Restraint: “The less conversation the better”**  *Well, there is a sort of a feeling of not being able to be heard, of being locked into permanent restriction or permanent amounts of medication, and the feeling of sort of a drop off point when you are going to be discharged and whether you are going to be able to handle the medication once you are out.*  **Abandonment**  *I went from being really really upset, to crying, to being fearful. It probably took me over 2 hours to get over the process of being undressed. That was terrifying to me—* *such personal transgression, put upon, being ordered to—it was terrifying and humiliating for it to be witnessed.* |
| Tomlin et al 2019  UK | To explore how patients experience forensic care | Mixed methods, constructivist, semi structured interviews, focus groups, thematic analysis | During admission | Forensic inpatient wards | Five participants in focus groups  13 individual interviews  Participants had a range of diagnoses and were from low, medium and high secure services | **Antecedent conditions to restrictive phenomena**  *I’m a section 3 so there’s no way I’m gonna wear handcuffs to go to the dentist.*  **Symbolic**  *“You need to give us this, this, this and this explain[ation]” but then if you can’t give them that explanation then “you’re being avoidant”. “You’re refusing to engage or in denial”. There’s always a reason why you’re not doing what you’re doing.*  **Soft, distal, and bureaucratic**  *It seems like everything is like a game like, you know what I mean like the ministry, the MoJ, there’s a delay there, there’s a delay there, there’s a delay here, like you say [one says] it’s waiting for bureaucracy to pull their finger out and the bureaucracy is too much.*  **Visible, routine, and coercive**  *…patronizing, it* [blanket restrictions] *feels kind of like untrusting*.  **Experiences of restrictive phenomena**  No quotes presented.  **Severity**  *Well it* [not being able to have perfume or a smartphone] *makes you feel different, you know, you’re sort of singled out different from the norm you know, from people on the outside, yes.*  **Salience**  *If they just say you can only have three sharp things in your room even though, some, them three sharp things might be sharper than other things you can’t have in your room, it does not make friggin’ sense at all.*  **Consequences of restrictiveness**  *I’ve been in and out of so many different places in my case in the last two or three years that erm, it’s become excessively difficult for me to actually function in my home.* |
| van Daalen-Smith et al 2020  Canada | To explore women’s experiences of psychiatric hospitalisation in Canada | Narrative methodology, Foucauldian analysis framework, semi structured interviews and focus groups, thematic analysis | Following admission, time frame of between two and 30 years between admission and interview | In the community | 12 women with psychiatric inpatient experience  Age range 22-62  Inpatient experiences were between two and 30 years prior to interview | **Descriptions of their experiences**  *You can’t imagine how bad it is until you experience it yourself.*  *It was worse than a prison… at least there you know how long you’ll be in for.*  **What was not helpful?**  *...because of the paradigm of broken brains, there’s no respect the point is missed. The point that...when people go through a crisis, there’s a reason for that crisis and the manifestations of that crisis have meaning...is missed, and that’s tragic.*  **Impact**  *It was the first time in my life that I considered suicide. They did that to me. It was from how I was treated in psychiatry.*  *I am now a broken person.*  **Keeping people quite and calm: making the docile patient**  No quotes presented.  **Harm: Ending off worse than pre-hospitalization**  *…the light inside you closes.*  **Betrayal**  No quotes presented.  **Indifference**  *No one really gets to know you.*  **Resistance**  *My mom told me to be a ‘good girl. ’Do what they say, and you’ll get out faster. She was right!* |
| Veale et al 2019  UK | To understand the patient experience of being a psychiatric inpatient at night | Semi structured interviews, content analysis | During admission | On the ward | 12 patients (four male and eight female)  Age range 21-64 | **Environmental disturbances: Sleep disruption due to observation procedures**  *They should have soft closing for the doors so that they stop slamming. The doors make me jump... with the loud banging... and this worsens my anxiety.*  **Environmental disturbances: Staff behaviour as a cause of sleep disturbance**  *I mean the light was irrelevant because by the time they clanked the keys and banged up and down other doors in the corridor I was wide awake...*  **Spending the night of a psychiatric ward: Other patients “invading” private space**  *One patient ... came in my room and at one point held me hostage and the nurses had to get me out.*  **Spending the night of a psychiatric ward: Uncomfortable with the presence of staff in the room at night**  *…* *when they come in and I see a silhouette it really frightens me.*  **Spending the night of a psychiatric ward: Suspension of privacy**  *I didn't like all the patients staring at me which they tended to do to have a good look through the window.*  **Well-being: Impact of sleep deprivation on emotional state**  *…* *staff kept coming every 15min ... startling me and waking me up at night.*  **Well-being: Poor communication with staff**  *There would be no interaction, even if I was awake, lying on the bed, they wouldn’t talk to me... Very very very little interaction if any at all. If you went to seek interaction, most did not give me the time.* |
| Wagstaff and Solts 2003  UK | To explore patients’ perspectives of ward rounds | Semi structured interviews, content analysis | During admission. Interview took place within one week of their last ward round | On the ward | Eight patients (three male and five female)  Age range 18-70 | **Decision-making**  *Prior to you coming in, they’ve already made an assessment about how they’re going to conduct the ward round.*  **Communication**  *If they’re all making a decision on my treatment, I’d expect that they’d all come and talk to me.*  **The number of people present**  *There were just too many people, I just wanted to talk to one person.*  **Practical arrangements**  *You don’t have a set time... if you go and see a doctor or a nurse you always have a set time... I think it’s very unprofessional... they just assume you’ll be sitting round.* |
| Wheatley et al 2013  UK | To explore the experience of transitioning from secure adolescent services to adult mental health services | Semi structured interviews, content analysis | During admission, transitioned from adolescent to adult services within the prior three months | On the ward | Eight patients who had transferred from adolescent medium secure services to adult secure services | **The aggressive behaviour of other patients**  *Some patients were really aggressive.*  *I was scared of being attacked.*  **Support from fellow service users and from staff**  *Staff didn’t comfort me.*  **Communication and information giving**  *I didn’t know what to expect on the day.*  *I wasn’t given information.*  *I didn’t like that I was told on the day of moving, just one hour before.* |
| Wilson et al 2017  UK | To explore the experience of physical restraint for patients and staff | Semi structured interviews, realist epistemological framework, thematic analysis | 10 were inpatient at time of interview. Three had been discharged at time of interview | On trust premises and in the wider community | 13 patients (six male and seven female)  Age range 18-65  Three had witnessed restraint, and 10 had been restrained themselves | **Is restraint a necessary evil?**  *I don’t think you’re ever going to negate it completely simply by the nature of people’s illness.*  **Emotional outcomes: Distress**  *I think it definitely scarred me...yes, distressing. Absolutely!*  **Emotional outcomes: Fear**  *Absolute terror! I was really scared...it was like something out of a horror movie...I was so terrified, I wet myself. I’ve never had such a terrifying experience in my life.*  **Emotional outcomes: Dehumanizing**  *That situation got out of control because they (other patient) weren’t talked to with compassion like a decent human being...people aren’t treated as ordinary flesh-and-blood human beings.*  **Relational outcomes: Power dynamics**  *I did feel that they have the power to do this to me...it’s a demonstration, even if unconsciously so, of what we can do to patients at this point.*  **Relational outcomes: Quality of patient-staff relationships**  *It left me with...a total distrust...a total vote of no confidence and no faith in anything they did, wanting to have absolutely nothing to do with any of them...all the time this is in my mind how they’ve treated me and how they treat other people, and obviously that affects relationships with them.*  **Restraint as a safety measure versus restraint as a cause of pain/injury**  *I was quite physically hurt on a couple of occasions.*  **Always a last resort?**  *Sometimes I think it’s necessary, because I’m doing quite a lot of harm to myself, but other times I don’t think it’s necessary...when I head bang...I don’t think I’m doing a lot of damage, but they still restrain me...if I’m just shouting, then I don’t think it’s necessary, because...not everybody gets restrained for shouting.*  **Role of communication**  *I also witnessed...a man who was obviously getting agitated...he’d been up all night...no one talked to him. By the end of the afternoon he was pulling televisions off the wall, and then of course...they raised their alarm and all piled in... there was no professional talking to him to settle him and distract him...and the poor man had to lose control.* |
| Wright et al 2015  UK | To explore the nature of service user involvement in the admission and discharge process of acute inpatient mental health care | Focus groups, thematic analysis | During admission | On the ward | 52 participants in total engaged in focus groups of ward staff, community staff and service users | **The lost voice of the service user**  *I was pulled in for what I thought was routine psychiatric appointment with Dr X and I was told ‘I want to send you home today’. Out of nowhere...so I didn’t take it well. I didn’t feel ready to go out...He said he was going to be honest because I deserved it. He had pressure from above to free the beds up and I said to him ‘so you don’t think I am well enough to go home but it’s just you need a few beds’ and so I was not very happy.* |
| Wyder et al 2015a  Australia | To describe the experience of people involuntarily admitted to acute mental health inpatient units | Semi structured interviews, inductive analysis approach | During admission, when the participants were ready for discharge | On the ward | 25 patients (14 female and 11 male)  Age range 24-65  Range of diagnoses and number of admissions | **Overall experience of the ITO [involuntary treatment order]**  No quotes presented.  **Staff potential to impact on ITO and hospital experiences**  *I get different injections and I try to explain that they are giving me too many, but I get dismissed and they don’t listen. It makes me feel upset and like a guinea pig.*  **What are good relationships? Finding time despite the busyness of the ward**  *Some nurses are always busy and they monitor us, and I don’t know what they do. They are constantly checking onus even at night and it often wakes me up. I am not sure what they are doing.*  **What are good relationships? Provision of information about the ward rules**  *I had no control and was given the pills and told just take this. I did not know what was going on. I needed structure in my day. I was left in the dark.*  **What are good relationships? Provision of information about ITO conditions**  *The only problem I found was when I asked about when the ITO was going to be lifted; I got shut down straightaway. Cause the nurses, I don’t know if they don’t know, but they don’t give you the time of day when you ask what’s happening with this . . . I didn’t even know I was going to get discharged ’til the day before, and I didn’t know that my ITO had been lifted. . . Just being informed a bit more would be better.* |
| Wyder et al 2015b  Australia | To describe the experiences of patients admitted involuntarily to a mental health unit, focusing on the legal processes | Semi structured interviews, interpretivist approach, inductive analysis | During admission, when the participants were ready for discharge | On the ward | 25 patients (14 female and 11 male)  Age range 24-65  Range of diagnoses and number of admissions | **ITO [involuntary treatment order] was experienced as an intrusion into their liberty and physical integrity**  *I just don't understand why I have to be here, that's all. I have done nothing wrong, why do I have to be on the frickin order?*  *I know the ITO is there to protect me but it impacts on the way I feel. It is scary, you have no rights. [...] Being involuntary means that I have no rights to say or do anything nor to make any legal decisions [...] I am part of the government now and they look after me.* |
| Wynn 2004  Norway | To explore the patients’ lived experience of restraint | Semi structured interviews, grounded theory | During admission, on average 11 days following episode of restraint | On the ward | 12 patients (nine male and three female)  Age range 21-60  Range of diagnoses  All had been restrained one+ times, both physically and chemically | **Patients’ perceptions of why physical and pharmacological restraint had been used**  *I could never have imagined this happening to me.*  **Patients’ opinions about if/how physical and pharmacological restraint could have been avoided**  *if they had asked ‘‘what is this?’’ then I could have responded that I was terribly anxious and needed to get the aggression out of my system . . . then we could get into a dialogue that could make the use of restraint completely unnecessary.*  **Patients’ experiences of the use of physical and pharmacological restraint**  *‘The worst part was not being able to move my body . . . I was completely helpless.*  **Patients’ thoughts about the consequences of having been restrained**  *It was completely unnecessary . . . it was an abusive act.* |
